# A Delphi Process for Reaching Nationwide Consensus on Antimicrobial Intravenous-to-oral Switch Criteria and Development of an Antimicrobial Intravenous-to-oral Switch Decision Aid

**DOI:** 10.1101/2022.11.12.22282255

**Authors:** Eleanor J Harvey, Kieran Hand, Dale Weston, Diane Ashiru-Oredope

## Abstract

**Introduction:** Antimicrobial stewardship (AMS) strategies, such as intravenous-to-oral switch (IVOS), promote optimal antimicrobial use, contributing to safer and more effective patient care and tackling antimicrobial resistance (AMR).

**Aim:** This study aimed to achieve nationwide multidisciplinary expert consensus on antimicrobial IVOS criteria for timely switch in hospitalised adult patients and to design an IVOS decision aid to operationalise agreed IVOS criteria in the hospital setting.

**Method:** A 4-step Delphi process was chosen to achieve expert consensus on IVOS criteria and decision aid; it included Step One) Pilot/1^st^ round questionnaire, Step Two) Virtual meeting, Step Three) 2^nd^ round questionnaire and Step 4) Workshop. This study follows the Appraisal of Guidelines for Research and Evaluation II instrument checklist.

**Results:** Step One questionnaire of 42 IVOS criteria had 24 respondents, 15 of whom participated in Step Two, where 37 criteria were accepted for the next step. Step Three had 242 respondents (England n=195, Northern Ireland n=18, Scotland n=18, Wales n=11); 27 criteria were accepted. Step Four had 48 survey respondents and 33 workshop participants; where consensus was achieved for 24 criteria and comments received on a proposed IVOS decision aid. Research recommendations include the use of evidence-based standardised IVOS criteria.

**Discussion and Conclusion:** This study achieved nationwide expert consensus on antimicrobial IVOS criteria for timely switch in the hospitalised adult population. For criteria operationalisation, an IVOS decision aid was developed. Further research is required to provide clinical validation of the consensus IVOS criteria and to expand this work into the paediatric and international settings.

## 1 Introduction

Antimicrobial stewardship (AMS) promotes optimal antimicrobial use, contributing to safer and more effective patient care and tackling antimicrobial resistance (AMR).^1-3^ AMR national action plans across the world^4^ set out to raise awareness on ‘when and how to use antimicrobials’,^5, 6^ implement AMS strategies specific to care settings,^7^ and reduce intravenous antimicrobial consumption.^8^

In secondary care, promoting timely antimicrobial intravenous-to-oral switch (IVOS) is typically considered a persuasive AMS strategy, whereby the use of standardised guidance or prompts improve stewardship. Persuasive AMS strategies have shown similar effectiveness in achieving sustainable AMS as restrictive strategies, such as the restriction of certain antibiotics.^9^

Equipping healthcare professionals with IVOS criteria and the rationale for their use in the clinical setting has been associated with a 48.7% increase in clinically appropriate switch decisions^10^ and a 14% reduction in intravenous therapy days.^11^ Studies highlight several IVOS benefits, including decreased risk of bloodstream and catheter-related infections, reduced medical equipment costs, carbon footprint and hospital length-of-stay, increased patient mobility and comfort, and released nursing time to care for patients.^12-15^ Timely IVOS has been shown to be safe and of equal efficacy to prolonged intravenous antimicrobial courses,^13, 16, 17^ with no negative impact on patient outcomes.^18^□

Hospital care settings have developed individualised IVOS policies,^19^ and within one country alone, criteria for timely IVOS varies considerably and lacks national consensus for standardisation.^20^

The aim of this initiative was to achieve nationwide consensus on antimicrobial IVOS criteria for safe and effective timely switch in hospitalised adult patients (over 18 years of age) receiving intravenous antimicrobial therapy. A secondary objective was to design a decision aid to operationalise agreed IVOS criteria for use in the hospital care setting. Target users for the criteria and decision aid are healthcare professionals involved in the treatment and management of infections in the specified patient population; including nursing or pharmacy teams who can prompt for an IVOS review and prescribers or infection specialists (all professions) who can make final decision for and implement switch.

## 2 Method

### 2.1 Ethical approval

This study did not require ethical approval according to the NHS Health Research Authority.^21^ It is a service evaluation and further development for the Start Smart – Then Focus antimicrobial stewardship toolkit.^22^ All data collected through questionnaires and virtual group gatherings were anonymised and obtained with informed consent.

### 2.2 Protocol and registration

This study follows the Appraisal of Guidelines for Research and Evaluation II instrument guidance^23^ and checklist (see **Supplementary Information**). The literature review informing the Delphi process has PROSPERO registration [CRD42022320343].^24^

### 2.3 Study question

The study question to answer was: ‘What are the minimum criteria necessary to achieve safe and effective antimicrobial IVOS in the hospital adult inpatient population?’.

### 2.4 Study design

A 4-step Delphi process was chosen to achieve nationwide consensus on IVOS criteria. The IVOS criteria introduced at Step One of the Delphi process were extracted from a combination of literature evidence, individual hospital IVOS policies and expert advice (see **Supplementary Information**).^20^

The Delphi process is a widely accepted technique to achieve group consensus,^25^ allows for participant anonymity and consideration of a breadth of expertise across wide geographical locations.^26^

### 2.5 Delphi process

**Figure 1** provides an overview of the study, including the Delphi process, the focus of this report, and the 4 steps of stakeholder involvement.

**Figure 1.**
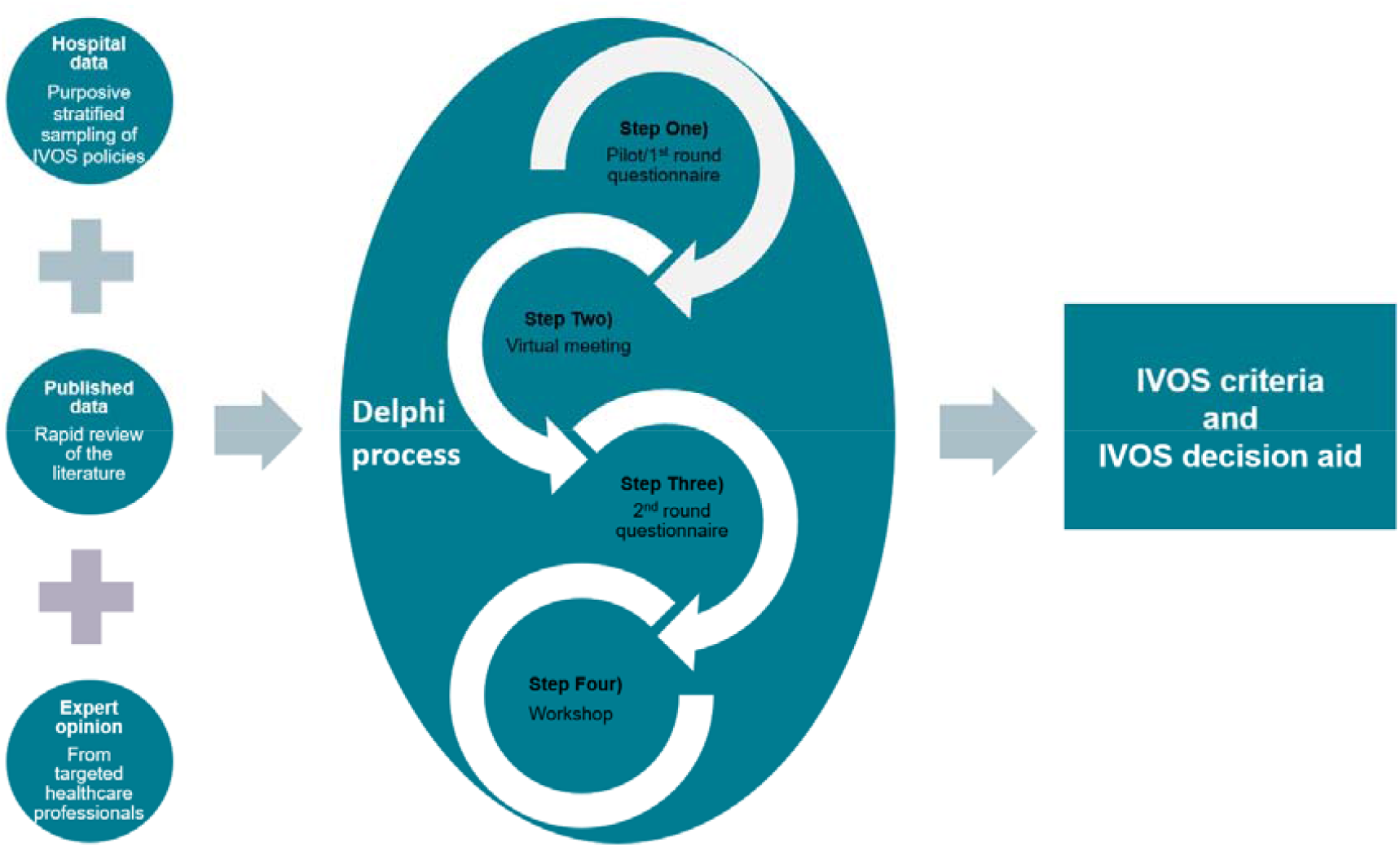
Overview of study undertaken to achieve UK-wide consensus of IVOS criteria and IVOS decision aid.

#### 2.5.1 Step One) Pilot/1st round questionnaire

Initial IVOS criteria identified from the published literature,^20^ a purposive sample of UK hospitals and expert opinion, were formatted into questions with 5-point Likert scale responses using Microsoft Forms (see **Supplementary Information**). Additional answer options included ‘Not applicable’ and ‘I don’t know’ The collated IVOS criteria were grouped into 5 sections: 1) *Timing of intravenous (IV) antimicrobial review*, 2) *Clinical signs and symptoms*, 3) *Infection markers*, 4) *Enteral route*, and 5) *Infection exceptions*.

Each section appeared twice in the questionnaire, for respondents to firstly consider the clinical relevance of criteria for safe and effective IV switch (a) Is each criterion clinically significant and does each criterion need to be met for a safe and effective IV switch? and b) Does each criterion apply to hospitalised adults prescribed IV antimicrobials?) and secondly to consider the ease with which criteria can be assessed in the clinical setting (How achievable is it to assess each criterion in practice?).

The questionnaire was sent by email to members of the English Surveillance Programme for Antimicrobial Utilisation and Resistance Oversight Group (ESPAUR OG), clinical fellows of the national oversight group led by UK Health Security Agency with members from more than 22 national health organisations and healthcare professionals including doctors, pharmacists and healthcare scientists, professional bodies across the four UK nations,,^27^ regional AMS leads, as well as select experts within the clinical practice setting to ensure representation across healthcare professions. Experts were selected based on their experience in AMS initiatives and occupation and included colleagues of the researchers or formal acquaintances through conferences.

Questionnaire responses were exported from Microsoft Forms to Excel for data analysis. Median scores and percentage agreement for all IVOS criteria were calculated. Consensus rules were adapted from Akhloufi H *et al*,^28^ and applied to answers regarding clinical relevance (ease of assessment answers served to inform, not dictate, consensus).

Consensus rules:

- Criteria with a clinical relevance median score of 4 (relevant) or 5 (very relevant) where there was agreement were accepted. Agreement was defined as ≥60% of participants assigning a score of 4 or 5.
- Criteria with a clinical relevance median score of 4 or 5 where there was lack of agreement were marked as uncertain and carried forward for consensus discussion in Step Two) Virtual meeting. Lack of agreement was defined as <60% of participants assigning a score of 4 or 5.
- Criteria with a clinical relevant median score <4 were rejected yet carried forward for consensus discussion in Step Two) Virtual meeting with the possibility for reinstating.
- Criteria with sufficient questionnaire comments to support a rephrase were rephrased and carried forward for consensus discussion in Step Two) Virtual meeting.

Options for IVOS criteria consensus therefore included: ‘accepted’, ‘rejected’, ‘rephrased’ or ‘uncertain’.

#### 2.5.2 Step Two) Virtual meeting

Respondents from Step One) Pilot/1^st^ round questionnaire were invited to a virtual meeting held using Microsoft Teams. The meeting was recorded with transcription, and a study researcher reviewed the transcription post meeting alongside comments typed in the Microsoft Teams channel, ensuring all opinions were comprehensively captured. The resulting proposed recommendations to improve questionnaire format and any IVOS criteria amendments were circulated to participants via email correspondence for approval and to provide an opportunity for further comments prior to Step Three. Options for IVOS criteria consensus were ‘accepted’, ‘new criteria proposed’, ‘rejected’ or ‘rephrased’.

#### 2.5.3 Step Three) 2^nd^ round questionnaire

Step Three) 2^nd^ round questionnaire (see **Supplementary Information**) was developed based on recommendations made during Step Two) Virtual meeting participants based on the Step One) Pilot/1^st^ round questionnaire. The questionnaire was cascaded via email to ESPAUR OG members, clinical fellows, Antimicrobial Pharmacist Network Wales members, hospital healthcare professionals within established AMS networks in England, Northern Ireland, Scotland and Wales and shared via social media (researchers’ Twitter accounts, using hashtag #IVOST and AMS WhatsApp groups).

The target number of respondents was 140 healthcare professionals representing multiple disciplines from across the UK. This is 0.01% of UK doctors, nurses and pharmacists (n=1,042,541).^29^

Questionnaire responses from Step Three included identifiable data of those participants who wanted ongoing involvement in AMS research. Responses were exported from Microsoft Forms to Excel and participant-identifiable information removed by one researcher.

Another researcher conducted data analysis on the anonymised data. Median scores and percentage agreement for all IVOS criteria were calculated. Consensus rules followed those of Step One; however, for criteria which differed with minor variance in wording, such as criteria in section 1) *Timing of IV antimicrobial review*, the criterion with highest median and/or percentage agreement was accepted. Options for IVOS criteria consensus were ‘accepted’ or ‘rejected’.

#### 2.5.4 Step 4) IVOS decision aid workshop

This step comprised of two stages: a pre-workshop questionnaire and a workshop discussion to finalise IVOS criteria and design a sample IVOS decision aid. The pre-workshop questionnaire was sent out with a draft IVOS decision aid (see **Supplementary Information**).

Survey responses were exported from Microsoft Forms to Excel, participant-identifiable information was removed by one researcher and another researcher conducted data analysis on the anonymised data. Proposed final amendments to IVOS criteria and feedback on the draft decision aid were extracted from the questionnaire responses, presented and discussed at the workshop to arrive at final consensus.

## 3 Results

### 3.1 Delphi process

#### 3.1.1 Step One) Pilot/1^st^ round questionnaire

The questionnaire was launched on 2 March 2022 with a closing date for completion of 10 March 2022 (extended until 15 March 2022). Twenty-four respondents completed the questionnaire. Most respondents were female (n=17, 71%) and based in England (n=23, 96%), with one respondent from Northern Ireland. Within England, there was representation from all regions, with highest representation from the Midlands (n=6, 25%) and London (n=5, 22%). Ethnicity data were not collected. Pharmacists as a professional group made up the majority of respondents (n=13, 54%), followed by Microbiologists/Infectious Diseases Physicians (n=5, 21%) (see **Table 1**).

**Table 1.** Professions of respondents who participated in the Delphi Step One) Pilot/1^st^ round questionnaire.

| Profession | Count |
| --- | --- |
| General Pharmacist | 7 |
| Specialist Microbiology/Infectious Diseases Pharmacist | 6 |
| Microbiologist/Infectious Diseases Physician | 5 |
| Microbiology/Infectious Diseases Nurse | 2 |
| General Physician | 1 |
| Surgeon | 1 |
| General Nurse | 1 |
| Healthcare Scientist | 1 |
| **Total** | **24** |

##### 3.1.1.1 IVOS criteria

Forty-two IVOS criteria were formatted into questions to determine clinical relevance and ease of assessment. Twenty-two criteria (52%) were accepted, ten (24%) were rejected, 9 (21%) were classified as uncertain, and one criterion (2%) was rephrased (see **Supplementary Information**). Criteria that were not accepted went forward for discussion in Step Two.

In section 1) *Timing of IV antimicrobial review*, the 4 criteria which differed with minor variance in wording were all accepted as all had median scores of 4 or 5 with ≥60% agreement in relevance for safe and effective IVOS. These criteria also went forward for discussion in Step Two to inform ongoing consensus for one timeframe for IV antimicrobial review.

The criterion that was rephrased was ‘abscess’ in section 5) *Infection exceptions*. Respondents’ comments included: ‘Abscess needs to be more clearly defined. Intra-abdominal abscess is different from a skin abscess or a tooth abscess’ and ‘The question on abscess is too broad’. The researchers suggested a rephrase to ‘Undrained abscess’ and agreed to carry forward for consensus discussion in Step Two.

#### 3.1.2 Step Two) Virtual meeting

The virtual meeting was held on 29 March 2022 for 70 minutes. There were 15 participants of the 24 respondents of Step One. Post meeting, proposed improvements to questionnaire format and proposed IVOS criteria amendments were emailed to participants on 31 March 2022, with an 8-day deadline for responses. Ten participants responded with further comments. Subsequently, the synthesised results of the Step Two) Virtual meeting are presented.

##### 3.1.2.1 Questionnaire format

Overall, consensus was reached to simplify questionnaire questions to ensure respondents would understand and interpret them correctly. Thus, Likert scales for clinical relevance and ease of assessment were converted into a Likert scale of agreement, such that, for example, ‘How clinically relevant is it to review IV antimicrobials within each specified timeframe’ and ‘How easy is it for the clinical team to review IV antimicrobials within each specified timeframe?’ was exchanged for ‘To what extent do you agree with the following timeframes of when intravenous-to-oral switch (IVOS) should be considered?’. The Likert scale responses were Strongly agree, Agree, Neither agree nor disagree, Disagree, Strongly disagree, Not applicable and I don’t know.

##### 3.1.2.2 IVOS criteria

Consensus was reached for 37 criteria; twenty-two (59%) were following rephrasing and 4 (11%) were new criteria proposed during Step Two. In section 1) *Timing of IV antimicrobial review*, all original criteria were reworded. Some were reworded to improve criterion clarity, such as ‘Review antimicrobial within 48 hours’ was amended to ‘IVOS should be considered within 48 hours of the first dose of IV antimicrobial being administered. Others were reworded both to improve criterion clarity and to inform whether consensus was partial towards ‘within’ or ‘after’ a set review timeframe, such as ‘Review antimicrobial within 48-72 hours’ was amended to ‘IVOS should be considered 48 hours after the first dose of IV antimicrobial is administered’. New criteria proposed included how often to undertake IVOS review, such as ‘Review daily thereafter if no switch within the first 48 hours’.

In section 3) *Infection markers*, heart rate, blood pressure and respiratory rate criteria were merged into an Early Warning Score (EWS) criterion. After criteria regarding White Cell Count (WCC) and C-Reactive Protein (CRP) were rejected at Step One, they were reinstated at Step Two as criteria with amended wording (see **Supplementary Information** for full list of criteria outcomes). Section 5) *Infection exceptions* title was changed to *Infection exclusions*.

#### 3.1.3 Step Three) 2^nd^ round questionnaire

The questionnaire was launched on 31 May 2022 with closing date for completion of 14 June 2022 (extended to 17 June 2022 to allow for further responses from the Northwest of England and Scotland and from nurses).

Two hundred forty-two respondents completed the questionnaire. This was 173% of the target number of respondents. Sixty-four per cent (n=154) were female, 33% (n=81) male and 3% (n=7) preferred not to state sex. Most respondents (n=165, 68%) were White, 27% (n=65) were from Asian/Asian British, Black/Black British, Mixed or other ethnic groups, and 5% (n=12) preferred not to say. Respondents were from across the UK; England (n=195, 81%), Northern Ireland (n=18, 7%), Scotland (n=18, 7%) and Wales (n=11, 5%). **Figure 2** shows count of respondents per region.

**Figure 2.**
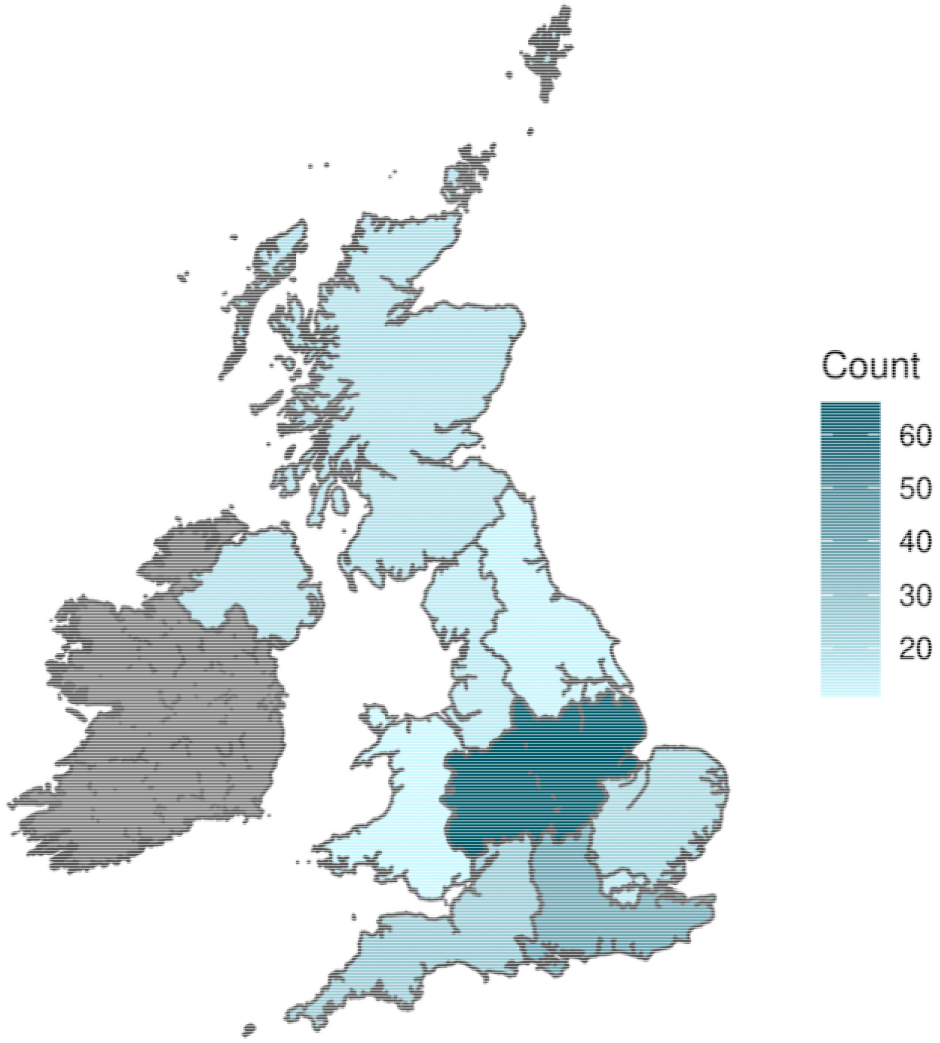
Map of Delphi Step Three) 2^nd^ round questionnaire respondents per region.

Microbiology or Infection Specialist Pharmacists comprised the largest group of respondents (n=65, 27%), followed by hospital General Physicians (n=55, 23%) (see **Table 2**). Sixty-five per cent of respondents (n=157) had over 10 years of experience within their profession. Of the specialists in infection, 18% (n=43) had over 10 years of experience in their specialist role.

**Table 2.** Professions of respondents who participated in Delphi Step Three) 2^nd^ round questionnaire.

| Profession of respondents | Count |
| --- | --- |
| Pharmacist (Microbiology or Infection Specialist) | 65 |
| Hospital General Physician | 55 |
| Medical Microbiologist/Infection Diseases Doctor | 38 |
| Pharmacist (General or non-Infection Specialist) | 36 |
| Nurse/Midwife (General or non-Infection Specialist) | 22 |
| Surgeon | 8 |
| Nurse (Antimicrobial Stewardship) | 8 |
| Allied Health Professional | 3 |
| Nurse (Infection Prevention Control) | 3 |
| Physician Associate | 1 |
| Healthcare Scientist | 1 |
| Specialist Pharmacy Technician | 1 |
| Dentist or Dental Nurse | 1 |
| **Total** | **242** |

Ninety-nine percent of respondents (n=239) stated they worked in an NHS acute hospital as their main area of work. Eighty-eight per cent of respondents (n=213) were currently actively involved in the clinical decision making, advising or implementing of antimicrobial IVOS, 7% (n=18) had previously been involved and 5% (n=11) had not been involved.

Results from the questionnaire’s general AMS questions included the finding that 67% of respondents (n=161) considered timely antimicrobial IVOS in clinically stable patients to have a positive impact on patient outcomes. The most frequently encountered barrier to IVOS was lack of time to review patient for IVOS suitability (n=97, 40%), followed by lack of senior agreement to switch (n=95, 39%), no suitable oral option available (n=92, 38%) and lack of a decision support tool for IVOS (e.g., checklist) (n=64, 26%).

Overall, respondents agreed that it was feasible or very feasible for both nursing and pharmacy teams to prompt medical colleagues to consider IVOS for eligible patients (see **Table 3**). There was a relative lack of agreement among respondents about how feasible it would be for nurses to ascertain infection markers for IVOS and infection exclusions to IVOS. There was also relative lack of agreement over how feasible it would be for pharmacy teams to ascertain clinical signs and symptoms relevant to IVOS.

**Table 3.** Feasibility for nursing teams (scored ‘feasible’ or ‘very feasible’) to prompt for IVOS in relation to each of the 5 sections of IVOS criteria.

| **Section** | **Feasible for nursing team to prompt for IVOS (%)** | **Feasible for pharmacy team to prompt for IVOS (%)** |
| --- | --- | --- |
| 1. Timing of IV antimicrobial review | 75.6 | 92.1 |
| 2. Clinical signs and symptoms | 69.5 | 54.1 |
| 3. Infection markers | 34.3 | 73.1 |
| 4. Enteral route | 81.4 | 75.3 |
| 5. Infection exclusions | 30.2 | 67.8 |

Fifty-two per cent of respondents (n=125) stated that electronic prescribing and medicines administration (ePMA) was not used within their place of work, whilst 43% (n=104) stated they did use ePMA. Five per cent (n=13) stated that ePMA was not applicable to their place of work, suggesting these respondents did not work in prescribing settings. Of the 43% of respondents (n=104) who did use ePMA, 43% (n=45) stated that ePMA facilitated antimicrobial IVOS, 25% (n=26) that it did not affect IVOS and 22% (n=23) that it hindered IVOS. Ten per cent (n=10) of respondents did not know how ePMA affected IVOS.

##### 3.1.3.1 IVOS criteria

Consensus was reached for 27 IVOS criteria. Regarding section 1) *Timing of IV antimicrobial review*, all timings had the same median score of 4, however the criterion ‘IVOS should be considered within 48 hours of the first dose of IV antimicrobial being administered’ had the highest percentage participant agreement of 78%, thus was accepted over the other options for timing criteria. Where no IVOS is conducted within the first 48 hours, reviewing ‘daily thereafter’, with a score of 88% agreement was accepted over reviewing ‘every two days thereafter’, which scored 6%.

Section 2) *Clinical signs and symptoms* had clear acceptance for the criterion ‘Clinical signs and symptoms are improving’, with median score of 5 and 95% participant agreement, compared to ‘Clinical signs and symptoms are not worsening’, with median score of 3 and 37% agreement. Section 3) *Infection markers* showed higher median and percentage participant agreement for the temperature criterion that included a timeframe: ‘Temperature is between 36-38°C for the past 24 hours’ was accepted with median score of 4 and 75% agreement, whilst ‘Temperature is between 36-38°C’ was rejected with median score of 3 and 48% agreement. For EWS, WCC and CRP criteria, ‘is improving’ wording was accepted over ‘is not worsening’ wording. All 8 criteria of section 4) *Enteral route* were accepted, as were all 12 criteria of section 5) *Infection exclusions* (see **Supplementary Information** for full list of criteria outcomes).

#### 3.1.4 Step Four) Workshop

A draft IVOS decision aid Version 0.0 (see **Supplementary Information**) was emailed out alongside a link to the pre-workshop questionnaire to the Step 3 respondents who had provided identifiable information (email addresses) to partake in ongoing AMS research. The email was sent on 5 August 2022, with closing date for completion of 12 August 2022 (extended until 15 August 2022). Forty-eight respondents completed the pre-workshop questionnaire. Fifty-four percent of respondents were female (n=26, 54%), with one person (2%) preferring not to state sex. Sixty-nine percent were of White ethnicity (n=33), followed by Asian or Asian British (n=7, 15%) and Black or Black British (n=3, 6%). Respondents were from across the UK; England (n=37, 77%), Scotland (n=8, 17%), Wales (n=2, 4%) and Northern Ireland (n=1, 2%). From within England, the Midlands region had most respondents (n=8, 22%) followed by East of England (n=7, 19%) and London (n=6, 16%).

The respondents included Pharmacists (n=21, 44%), Doctors (n=20, 42%), Nurses (n=6, 13%) and Physician Associates (n=1, 2%). Eighty-five percent of respondents (n=41) worked in an acute NHS hospital as their main area of work. Eighty-three per cent of respondents (n=40) were actively involved in the clinical decision making, advising or implementing of antimicrobial IVOS, 13% (n=6) had previously been involved and 4% (n=2) had not been involved.

The draft IVOS decision aid Version 0.0 was updated to Version 0.1 based on feedback received from questionnaire responses and feedback from the behavioural scientist study researcher. IVOS decision aid Version 0.1 was presented at the workshop. The workshop was held on 17 August 2022 for 90 minutes, facilitated by study researchers. There were 33 participants, including from England (n=28, 85%), Scotland (n=3, 9%), Northern Ireland (n=1, 3%) and Wales (n=1, 3%). Subsequently, the summarised feedback and discussion of the Step Four) Workshop are presented.

##### 3.1.4.1 IVOS criteria

UK-wide consensus was reached for 24 IVOS criteria. Criteria in section 3) *Infection markers* were rephrased, whereby EWS, WCC and CRP criteria qualifier wording changed from ‘is improving’ to ‘is decreasing’, ‘is trending towards the normal range’ and ‘is decreasing’, respectively. Criteria in section 4) *Enteral route* were merged to decrease overall number of criteria; for example, ‘Gastrointestinal tract must be functional’ and ‘No evidence of malabsorption’ were merged into ‘Gastrointestinal tract is functioning with no evidence of malabsorption’. The criterion ‘Patient can tolerate/swallow oral option’ was amended for clarity to ‘Safe swallow or enteral tube administration’.

Section 5) *Infection exclusions* title was further changed to *Special considerations* to avoid giving the impression that exclusions were absolute. The criterion ‘Bacteraemia, including *Staphylococcus aureus*’ was amended to ‘Bloodstream infection’ to improve accessibility of language (see **Supplementary Information** for criteria outcomes).

##### 3.1.4.2 Draft IVOS decision aid

Seventy-four comments were received through Step 4 questionnaire responses, 24 (32%) were particular to IVOS decision aid design which led to changes from Version 0.1 to Version 1 (see **Supplementary Information**).

For operationalisation of IVOS criteria, electronic prescribing system prompts were the preferred option (see **Table 4**).

**Table 4.** Survey respondents’ ideal formats to deploy the IVOS criteria for implementation in hospital settings.

| Format | Count (percentage) |
| --- | --- |
| Electronic prescribing system prompt | 39 (81) |
| Sticker to add to drug chart and/or medical notes | 18 (38) |
| Poster for e.g., treatment room | 12 (25) |
| Smartphone app | 12 (25) |
| Page to add to medical notes | 9 (19) |
| Other | 7 (15) |
| Pocket-sized card | 2 (0.4) |

#### 3.1.5 Final IVOS criteria

National consensus was reached for 24 IVOS criteria (see **Table 5**).

**Table 5.** Antimicrobial intravenous-to-oral (IVOS) criteria for timely switch categorised into 5 sections.

| Section | Antimicrobial IVOS criteria |
| --- | --- |
| 1. **Timing of IV antimicrobial review** | IVOS should be considered within 48 hours of the first dose of IV antimicrobial being administered<br>If no IVOS within first 48 hours, review daily thereafter |
| 2. **Clinical signs and symptoms** | Clinical signs and symptoms of infection are improving |
| 3. **Infection markers** | Temperature is between 36-38°C for the past 24 hours<br>Early Warning Score is decreasing<br>White Cell Count is trending towards the normal range*<br>C-Reactive Protein is decreasing* |
| 4. **Enteral route** | Gastrointestinal tract is functioning with no evidence of malabsorption<br>Safe swallow or enteral tube administration<br>Suitable oral switch option available, considering oral bioavailability, any clinically significant drug interactions or patient allergies<br>No significant concerns over patient adherence to oral treatment<br>No vomiting within the last 24 hours |
| 5. **Special considerations** | Deep-seated infection<br>Infection requiring high tissue concentration<br>Infection requiring prolonged IV therapy<br>Critical infection with high risk of mortality<br>*Specific infections for special consideration are:*<br>- Bloodstream infection<br>- Empyema<br>- Endocarditis<br>- Meningitis<br>- Osteomyelitis<br>- Severe or necrotising soft tissue infections<br>- Septic arthritis<br>- Undrained abscess |
| *To note: these infection markers could also indicate non-infectious inflammation or be affected by other factors e.g., steroid treatment. |  |

## 4 Discussion

This study reached nationwide consensus on 24 evidence-based antimicrobial IVOS criteria for timely switch in the hospitalised adult population. The researchers recommend the use of evidence-based IVOS criteria when designing IVOS decision aids for hospital settings to improve patient care. For IVOS criteria operationalisation, a sample IVOS decision aid is suggested; it was informed by behaviour science and designed to maximise ease and simplicity of use.

A Bayesian-like approach to antibiotic treatment decision-making highlights the role of different healthcare professionals (HCPs) in a patient’s care pathway.^30^ Research shows the benefit of multidisciplinary HCP involvement in IVOS,^3^ and this study corroborates the feasibility of a partnership approach, whereby nursing and pharmacy teams play a role in reviewing IVOS criteria and prompt prescribers to consider switch. An Australian study by Mostaghim M *et al*. captured that 75% of study participant nurses recognised their role in AMS but acknowledged limited knowledge regarding antimicrobials.^31^ An Italian study demonstrated that pharmacists contributing to IVOS decisions led to improvements in appropriate dosing and reduced medication costs.^32^ Across the hospital care setting, these professionals are underutilised within AMS strategies;^31^ efforts are needed to develop their understanding of IVOS and define the roles that will lead to their increased participation.

The consensus of study participants was that IV antimicrobial review should occur within 48 hours of initiation, which differs to prior national guidance stipulating review at 48-72 hours.^22^ An early review requires HCP collaboration, and could lead to earlier switch and increased IVOS benefits.^33^ The novel criterion suggested, ‘Early Warning Score is decreasing’, builds on the widespread international use of EWS in clinical practice to detect patient deterioration.^34^ This criterion amalgamates vital signs criteria of heart rate, blood pressure and respiratory rate that appeared with inconsistent definitions and thresholds across the literature and individual hospital IVOS policies. Infection exceptions or exclusions were accepted as special considerations, as study participants agreed that, for the listed criteria infections, there must be a clearly documented plan or specialist input but they did not necessarily preclude IVOS.^35, 36^

The potential benefits of timelier IVOS to the National Health Service in England are considerable, where 18 million standard treatment days (defined daily doses) of IV antibiotics were dispensed in 2021-22.^37^ Reducing the average duration of IV treatment courses by one day could result in (per annum) up to 2,500 fewer bloodstream infections, release 1.7 million hours of nursing time and reduce drug expenditure by £35 million.^37-40^ Research indicates that timely IVOS interventions for hospitalised patients can result in reduced length-of-stay; a one day reduction in length-of-stay for all patients treated with IV antimicrobials in English hospitals would release up to 5 million occupied beds to facilitate recovery of services following the COVID-19 pandemic.^37, 41, 42^

Despite these considerable benefits, the published literature identifies barriers to IVOS uptake in practice.^43^ This study reveals that lack of time to review patients for IVOS suitability is the most common barrier, followed by lack of senior agreement to switch. Clarity in IVOS intent and criteria for switch must be communicated effectively, to ensure patients are not exposed to the risk of IV treatment any longer than necessary.^44^ Electronic alert systems to support IVOS have been validated,^45^ and survey responses from this study corroborate the preference for IVOS electronic prescribing system prompts. As demonstrated by questionnaire responses, not all settings have electronic prescribing systems, thus alternative validated resources to promote IVOS should be developed.

Design informed by behaviour science for intervention development, such as IVOS decision aid development, contributes to behaviour change.^46^ Prescribing behaviour can be defined, for example, by social norms, evasion of responsibility or ‘decision inertia’,^47^ and can vary between care settings and contexts.^3^ This study provides a suggested IVOS decision aid; however, it is acknowledged that further incentives for behaviour change occur in the clinical care context, through education and training and collaboration, thus implementation should be supported by ongoing research.

### 4.1 Strengths and limitations

Study strengths include a large Delphi process with national stakeholder involvement, including representation from the four countries of the UK and multiple health professional groups. Additionally, criteria going into the Delphi process was informed by published literature, hospital IVOS policies in current use and expert opinion. Achieving national consensus allows IVOS criteria to be used for national incentive schemes to improve clinical practice. The UK-wide questionnaire exceeded target response rate, increasing validity of IVOS criteria consensus. However, it should be noted that the target response rate was an overestimate of the size of the stakeholder group as not all would work within the acute setting where IVOS is part of routine practice.

A study limitation was that a small minority of respondents were not actively working in clinical environments where IVOS was practised, however, many of these respondents had past experience of working in clinical settings. The final set of IVOS criteria was developed by expert consensus from a longer list of evidence-based criteria identified from the published literature, hospital policies and expert opinion; but this specific set of criteria has not been tested for clinical effectiveness and safety.

### 4.2 Further research

Assessment of antimicrobial IVOS criteria in acute hospital settings should be undertaken to test the feasibility and clinical safety of IVOS criteria and to test the effectiveness of any resources for implementation in practice and identify potential resource implications of adopting the recommendations. The standardisation of criteria allows for future auditing of NHS hospital IVOS practice. Future research should focus on the validation of IVOS criteria and expand use internationally. Paediatric clinical practice was out of scope and requires a separate programme of work with paediatric specialists. Updates to both IVOS criteria and the IVOS decision aid are recommended at least every 5 years, or sooner if relevant evidence is published, via a similar study, to ensure AMS interventions remain contemporary and relevant for safe and effective patient care.

### 4.3 Conclusion

In the UK hospital setting, expert consensus has been reached for nationwide IVOS criteria to prompt and support antimicrobial IVOS decisions for adult inpatients treated with IV antimicrobials.

## Supporting information

Delphi Step Four_Workshop_IVOS Decision Aid Versions

Delphi Step Four_Workshop_Pre-workshop Questionnaire

Delphi Step One_Pilot1st Round_Questionnaire

Delphi Step Three_2nd Round_Questionnaire

Delphi_AGREE Reporting Checklist

Delphi_All Steps_Criteria Outcomes

Pre Delphi_IVOS Criteria

## Data Availability

All data produced in the present study are available upon reasonable request to the authors

## 5 Author contributions

Conceptualisation: DAO, KH. Methodology: DAO, KH. EH. First paper draft: EH. All authors reviewed and approved final version.

## 6 Acknowledgements

We are grateful to all study participants across the 4-step Delphi process (see **Supplementary Information**). Additionally, we would like to thank Dr Alex Orlek for the map and count of UK-wide results.

## 9 Data availability statement

All data relevant to the study are included in the article or uploaded as supplemental information.

