## Supplementary material for "A Delphi Process for Reaching Nationwide Consensus on Antimicrobial Intravenous-to-oral Switch Criteria and Development of an Antimicrobial Intravenous-to-oral Switch Decision Aid": Delphi Step Four_Workshop_IVOS Decision Aid Versions

UK Health Security Agency

### Version 0.0: Antimicrobial Intravenous-to-Oral Switch (IVOS) Tool

#### Co-produced through a UK-wide consensus process involving 269 multi-disciplinary participants

DRAFT VERSION

Intravenous antimicrobial initiation – Date: _______________ Time: _______________
IVOS assessment – Date: _______________ Time: _______________

PROMPT FOR SWITCH (by nursing/pharmacy teams)

Verified by prescriber or infection specialist

| | | |
|---|---|---|
| **1.Timing of IV antimicrobial review** | | |
| 1.1 IVOS should be considered within 48 hours of the first dose of IV antimicrobial being administered | | ☐ |
| 1.2 If no IVOS within first 48 hours, assess daily thereafter | | ☐ |
| **2. Clinical signs and symptoms** | | |
| 2.1 Clinical signs and symptoms are improving | | ☐ |
| **3. Infection markers*** | | |
| 3.1.Temperature has been between 36-38°C for the past 24 hours | Temperature is _______ | ☐ |
| 3.2. Early Warning Score (EWS), e.g. NEWS2, is improving | EWS is _______ | ☐ |
| 3.3. White Cell Count (WCC) is improving towards normal | WCC is _______ | ☐ |
| 3.4. C-Reactive Protein (CRP) is improving towards normal | CRP is _______ | ☐ |
| **4. Enteral route** | | |
| 4.1. No significant concerns over patient adherence to oral treatment | | ☐ |
| 4.2. No vomiting within the last 24 hours | | ☐ |
| 4.3. Gastrointestinal tract must be functional with no evidence of malabsorption | | ☐ |
| 4.4. Suitable oral switch option available, with acceptable oral bioavailability | | ☐ |
| 4.5. Patient can swallow oral option or administer via enteral tube | | ☐ |
| 4.6. No contraindication or clinically significant drug interaction affecting oral switch option | | ☐ |
| 4.7. No clinically significant allergy to oral switch option | | ☐ |

ASSESS FOR SWITCH (by prescriber or infection specialist)

| | |
|---|---|
| **5. Infection exclusions** | |
| 5.1. Deep-seated infection | ☐ |
| 5.2. Infection requiring high tissue concentration of antimicrobial | ☐ |
| 5.3. Infection requiring prolonged IV therapy | ☐ |
| 5.4. Critical infection with high risk of mortality | ☐ |
| 5.5. Bacteraemia, including *Staphylococcus aureus* | ☐ |
| 5.6. Empyema | ☐ |
| 5.7. Endocarditis | ☐ |
| 5.8. Meningitis | ☐ |
| 5.9. Osteomyelitis | ☐ |
| 5.10. Severe or necrotising soft tissue infections | ☐ |
| 5.11. Septic arthritis | ☐ |
| 5.12. Undrained abscess | ☐ |

##### PROMPT FOR SWITCH : OUTCOME

Nurse/pharmacist to review criteria to prompt prescriber or infection specialist for IVOS switch

Criteria 1.1 – 4.2 all ☑ → Prompt prescriber or infection specialist to consider IV to oral switch

Criteria 1.1 – 4.2 any ☒ → Assess for IVOS the next day unless clear plan documented in notes

##### ASSESS FOR SWITCH : OUTCOME

Prescriber or infection specialist to assess for IVOS switch

Criteria [illegible].7 all ☐
Criteria 5.1 – 5.1[illegible] all ☐
→ Consider IV to oral switch

Draft Version 0.0
05/08/2022

*Note: infection markers also include markers which could indicate inflammation rather than infection

**Antimicrobial Intravenous-to-Oral Switch (IVOS) Criteria – Project information**

These criteria have been developed through:
- Collation of criteria from a sample of 45 acute hospital IVOS policies
- Completion of a rapid literature review to validate selection of IVOS criteria
- Three-step Delphi consensus-gathering process involving 269 participants to agree evidence-based, UK-wide IVOS criteria for hospitalised adult patients

The Delphi process consisted of:
- Step 1) Pilot/1st round questionnaire (24 respondents)
- Step 2) Virtual meeting to discuss findings of pilot/1st round questionnaire (15 participants)
- Step 3) 2nd round questionnaire (242 respondents)

The total of 269 participants across the 3-step process accounts for individuals who provided expert advice outside of questionnaires and virtual meeting, and ensures individuals who participated in more than one step are only included once in total count.
The additional Step 4) Workshop aims to finalise the agreed criteria as a tool for IVOS.

This project has been led by Eleanor Harvey and Prof Diane Ashiru-Oredope of the UK Health Security Agency, in collaboration with Dr Kieran Hand of NHS England. The IVOS tool was designed with input from Dr Dale Weston of the Behavioural Science and Insights Unit at the UK Health Security Agency.

UK Health Security Agency

### Version 0.1:
### National Antimicrobial Intravenous-to-Oral Switch (IVOS) Criteria for Early Switch (within 48 hours of IV antimicrobial initiation) (A6/8/10/12/14/17/26/38, B18, C8) and daily thereafter

PROMPT FOR SWITCH (by nursing/pharmacy teams)

DRAFT VERSION

| **1.Timing of IV antimicrobial review** | | |
|---|---|---|
| 1.1. Review IVOS should be considered (A7) within 48 hours of the first dose of IV antimicrobial being administered | | ☐ |
| 1.2. If no IVO[illegible] within first 48 hours, review daily thereafter | | ☐ |
| **2. Cli[illegible]al signs an[illegible] symptoms** | | |
| 2.1. [illegible]inical sig[illegible] and symptoms of infection (A30) are improving e.g. *patient feeling better* (A7) | | ☐ |
| **3. Infection m[illegible]ers** | | |
| 3.1. Temperature h[illegible] between 36-38°C for the past 24 hours | Temperature is _______ | ☐ |
| 3.2. Early Warnin[illegible] Score (EWS), e.g. NEWS2, is improving or stable (A30) | EWS is _______ | ☐ |
| 3.3. White Cell Co[illegible] (W[illegible]CC) is [illegible]mproving ~~towards normal~~ (A6) | WCC is _______ | ☐ |
| 3.4. C-Reactive Protein (C[illegible]) is improving ~~towards normal~~ (A6) | CRP is _______ | ☐ |
| **4. Enteral route** | | |
| 4.1. No significant concerns over pa[illegible]ent adherence to oral treatment | | ☐ |
| 4.2. No vomiting within the last 24 hours | | ☐ |
| 4.3. Gastrointestinal tract must be functional [illegible] no evidence of malabsorption | | ☐ |
| 4.4. Suitable oral switch option availabl[illegible] ~~with acc[illegible]ptable oral bioavailability~~ (A37) | | ☐ |
| 4.5. Patient can swallow oral option [illegible]r administer via enteral tube | | ☐ |
| 4.6. No contraindication or clinically significant drug in[illegible]raction affecting oral switch option | | ☐ |
| 4.7. No ~~clinically significant~~ allergy to oral switch op[illegible]n or other clinically significant adverse reaction (A6) | | ☐ |
| **5. Infection exclusions > Special considerati[illegible] ([illegible] any tha[illegible] apply)** | | |
| 5.1. Deep-seated infection | | ☐ |
| 5.2. Infection requiring high tissue concentration of a[illegible]icrobial t[illegible]rapy (A5) | | ☐ |
| 5.3. Infection requiring prolonged IV therapy | | ☐ |
| 5.4. Critical infection with high risk of mortality | | ☐ |
| 5.5. Gram positive (A11/12/13) b~~B~~loodstream infection~~, [illegible]cluding [illegible]aph[illegible]ococcus aureus bacteraemia~~ (A6) | | ☐ |
| 5.6. Empyema | | ☐ |
| 5.7. Endocarditis | | ☐ |
| 5.8. Meningitis | | ☐ |
| 5.9. Osteomyelitis | | ☐ |
| 5.10. Severe or necrotising soft tissue infections | | ☐ |
| 5.11. Septic arthritis | | ☐ |
| 5.12. Undrained abscess | | ☐ |

Verified by prescriber or infection specialist

ASSESS FOR SWITCH (by prescriber or infection specialist)

*Note: infection markers also include markers which could indicate inflammation rather than infection

#### PROMPT FOR SWITCH : OUTCOME

Nursing/pharmacy teams to assess criteria for prompting prescriber or infection specialist for IVOS switch

Criteria 1.1 – 4.2 all ☑
⇩
Prompt prescriber or infection specialist to consider IV to oral switch

Criteria 1.1 – 4.2 any ☒
⇩
Review for IVOS the next day unless clear plan documented in notes

#### ASSESS FOR SW[illegible]CH : [illegible]TCOME

Prescriber or infect[illegible] spec[illegible]alist to assess for IVOS sw[illegible]ch

Criteria 1.1 – 4.7 [illegible]
Criteria 5.1 – 5.12 ~~all~~ any ~~✗~~ ✓
⇩
Consider IV to oral switch

Draft Version 1.
05/08/2022

Intravenous antimicrobial initiation – Date: _______________ Time: _______________
IVOS assessment – Date: _______________ Time: _______________

© Crown copyright 2022

UK Health Security Agency

### Version 1: National Antimicrobial Intravenous-to-Oral Switch (IVOS) Sample Decision Aid for Timely Switch

#### IVOS criteria co-produced through a UK-wide consensus process involving 279 multi-disciplinary participants

**Why use this IVOS decision aid?**

IVOS is an important antimicrobial stewardship intervention.[1,2] The literature highlights several IVOS benefits, including decreased risk of bloodstream and catheter-related infections, reduced equipment costs, carbon footprint and hospital length-of-stay, increased patient mobility and comfort, and released nursing time to care for patients.[3,4]

**When to use this IVOS decision aid?**

The **audit standard** recommended for the implementation of this tool is that all patients on intravenous (IV) therapy should be reviewed **promptly from first dose** of IV antimicrobial with formal review completed **within 48 hours** and daily thereafter, unless clearly documented exemptions.

**Does your patient have any of the following infections for special consideration?**

*To note: An IVOS within 48 hours may still be indicated for deep-seated infections, infections requiring high tissue concentration or prolonged intravenous antimicrobial therapy, or critical infections with high risk of mortality, such as those listed below.*

| | |
|---|---|
| Bloodstream infection | Y/N |
| Empyema | Y/N |
| Endocarditis | Y/N |
| Meningitis | Y/N |
| Osteomyelitis | Y/N |
| Severe or necrotising soft tissue infections | Y/N |
| Septic arthritis | Y/N |
| Undrained abscess | Y/N |

*If **NO**, continue* — *If **YES**, check for clearly documented plan or seek specialist advice*

**1a. Enteral route**

| | |
|---|---|
| 1.1. Is the patient's gastrointestinal tract functioning, e.g. no evidence of malabsorption? | Y/N |
| 1.2. Is the patient's swallow or enteral tube administration safe? | Y/N |
| 1.3. Is there a suitable oral switch option available?<br>*Considering e.g. oral bioavailability, any clinically significant drug interactions or patient allergies* | Y/N |

*If **YES**, continue* — *If **NO**, reassess in 24 hours*

**1b. Enteral route continued**

| | |
|---|---|
| 1.4. Are there any significant concerns over patient adherence to oral switch option? | Y/N |
| 1.5. Has the patient vomited within the last 24 hours? | Y/N |

*If **NO**, continue* — *If **YES**, reassess in 24 hours*

**2. Clinical signs and symptoms**

| | |
|---|---|
| 2.1. Are the patient's clinical signs and symptoms of infection improving?<br>*E.g. patient feeling better, patient able to eat and drink* | Y/N |

**3. Infection markers**

| | | |
|---|---|---|
| 3.1. Has the patient's temperature been between 36-38°C for the past 24 hours? | Temperature: ____ | Y/N |
| 3.2. Is the patient's Early Warning Score (EWS) decreasing? | EWS: ____ | Y/N |
| 3.3. Is the patient's White Cell Count (WCC) trending towards the normal range?* | WCC: ____ | Y/N |
| 3.4. Is the patient's C-Reactive Protein (CRP) decreasing?* | CRP: ____ | Y/N |

*If **YES** to 2.1-3.4, prompt or assess for switch* — *If **NO** to 2.1-3.2*, reassess in 24 hours*

**PROMPT FOR SWITCH:**
Nursing/pharmacy teams to prompt prescriber or infection specialist to consider IV to oral switch.

**ASSESS FOR SWITCH:**
Prescriber or infection specialist to consider IV to oral switch.

Intravenous antimicrobial initiation: Date: __________ Time: __________ Name: ____________

IVOS first assessment (daily thereafter): Date: __________ Time: __________ Name: ____________

IVOS switch: Date: __________ Time: __________ Name: ____________

*To note: these infection markers could also indicate inflammation or be affected by e.g. steroid treatment, 'Prompt for switch' or 'Assess for switch' may still occur if they are the only markers not met.

Version 1. 28/09/2022


© Crown copyright 2022
