## Supplementary material for "A Delphi Process for Reaching Nationwide Consensus on Antimicrobial Intravenous-to-oral Switch Criteria and Development of an Antimicrobial Intravenous-to-oral Switch Decision Aid": Delphi Step Four_Workshop_Pre-workshop Questionnaire

### Antimicrobial Intravenous-to-oral Switch Criteria and Tool for Implementation

Thank you for your contribution to the development of evidence-based, UK-wide antimicrobial intravenous-to-oral switch (IVOS) criteria, and for agreeing to be contacted.

We invite you to provide feedback on the draft version of the IVOS tool.

This survey will take approximately 10 minutes to complete and will be open until COP **Friday 12 August 2022**. **(Extended to 15th August 2022)**
**Please ensure you click submit at the end of the form**, otherwise none of your responses will be recorded.
The information you provide will remain anonymous.

Please direct any queries to:
Eleanor Harvey – Chief Pharmaceutical Officer Clinical Fellow, UKHSA
Prof Diane Ashiru-Oredope – Chair, English Surveillance Programme for Antimicrobial Utilisation and Resistance and UKHSA's Lead Pharmacist for HCAI, AMR, AMU, Fungal and Sepsis Division

**About this study**

The intravenous-to-oral switch (IVOS) criteria have been developed through:
- Collation of criteria from a sample of 45 acute hospital IVOS policies
- Completion of a rapid literature review to evaluate IVOS criteria
- Three-step Delphi consensus-gathering process involving 269 participants to agree evidence-based, UK-wide IVOS criteria for hospitalised adult patients.

The Delphi process consisted of:
- Step 1) Pilot/1st round questionnaire (24 respondents)
- Step 2) Virtual meeting to discuss findings of the pilot/1st round questionnaire (15 participants)
- Step 3) 2nd round questionnaire (242 participants)

The total of 269 participants across the 3-step process accounts for individuals who provided expert advice outside of questionnaires and virtual meeting, and ensures individuals who participated in more than one step are only included once in total count.

The additional Step 4) Workshop aims to finalise the agreed criteria as a tool for IVOS.

This project has been led by Eleanor Harvey and Prof Diane Ashiru-Oredope of the UK Health Security Agency, in collaboration with Dr Kieran Hand of NHS England.

The IVOS tool has been designed with input from Dr Dale Weston of the Behavioural Science

#### Consent

Consent form for participants in the Delphi process to inform an evidence-based anti-microbial IVOS tool for hospitalised adult patients. The information you provide will remain anonymous.

1

I consent to participating in this survey and understand that any information I provide will be used to help inform an evidence-based antimicrobial IVOS tool for hospitalised adult patients. *

- ◯ Yes
- ◯ No

#### About you

2

Are you actively involved in the clinical decision making, advising or implementing of antimicrobial IVOS? *

- Yes
- No
- Not currently

3

What is your profession?
*If 'Other', please include profession.* *

- ○ Medical Microbiologist/Infection Diseases Doctor
- ○ General Physician
- ○ Surgeon
- ○ Dentist or Dental Nurse
- ○ Nurse (Antimicrobial Stewardship)
- ○ Nurse (Infection Prevention Control)
- ○ Nurse/Midwife (General or non-Infection Specialist)
- ○ Pharmacist (Antimicrobial or Infection Specialist)
- ○ Pharmacist (General or non-Infection Specialist)
- ○ Allied Health Professional
- ○ Clinical Researcher (Infection Specialist)
- ○ Clinical Researcher (General)
- ○ Healthcare Scientist
- ○ Other

4

If an infection specialist, for how long have you been in your specialist role? *

- ◯ 0-1 year
- ◯ 1-2 years
- ◯ 2-5 years
- ◯ 5-10 years
- ◯ 10+ years
- ◯ Not applicable

5

How many years have you been practising within your profession? *

[ ]

6

Which of the following describes your main area(s) of work? *

- ☐ NHS Non-teaching Acute Trust
- ☐ NHS Specialist Acute Trust
- ☐ NHS Teaching Acute Trust
- ☐ Independent Hospital
- ☐ Community Health Trust
- ☐ Mental Health Trust
- ☐ NHS England
- ☐ UKHSA
- ☐ Other Arms Length Body/Organisation
- ☐ Academia
- ☐ Government
- ☐ General Practice/CCG
- ☐ Other

7

Which country do you work in? *

- ◯ England
- ◯ Northern Ireland
- ◯ Scotland
- ◯ Wales
- ◯ Other

8

Which region do you work in? *

- ◯ East of England
- ◯ London
- ◯ Midlands
- ◯ North East and Yorkshire
- ◯ North West
- ◯ South East
- ◯ South West
- ◯ National

9

Please provide the first half of your organisation's post code (to ensure UK-wide representation).

#### The IVOS tool

10

Are you in agreement with the IVOS criteria included in each section of the tool? *

| | Yes | Somewhat | No | I don't know |
|---|---|---|---|---|
| 1. Timing of IV antimicrobial review | ○ | ○ | ○ | ○ |
| 2. Clinical signs and symptoms | ○ | ○ | ○ | ○ |
| 3. Infection markers | ○ | ○ | ○ | ○ |
| 4. Enteral route | ○ | ○ | ○ | ○ |
| 5. Infection exclusions | ○ | ○ | ○ | ○ |

11

If you responded 'Somewhat', 'No' or 'I don't know' to the previous question, please use the box below to provide further details as to why. Please clearly state which section you are referring to.

12

Are you in agreement that criteria 1.1-4.2 are suitable to be included as part of the 'prompt for switch' by nursing/pharmacy teams? *

- Yes
- Somewhat
- No
- I don't know

13

If you responded 'Somewhat', 'No' or 'I don't know' to the previous question, please use the box below to provide further details as to why.

14

Are you in agreement that the 'prompt for switch' and 'assess for switch' outcomes are accurate? *

| | Yes | Somewhat | No | I don't know |
|---|---|---|---|---|
| Prompt for switch: outcome | ○ | ○ | ○ | ○ |
| Assess for switch: outcome | ○ | ○ | ○ | ○ |

15

If you responded 'Somewhat', 'No' or 'I don't know' to the previous question, please use the box below to provide further details as to why. Please clearly state which outcome you are referring to.

16

In your opinion, what would be the top TWO ideal formats to deploy the IVOS criteria for implementation in hospital settings? *If 'Other', please state format* *

- ☐ ePrescribing system prompt
- ☐ Smartphone app
- ☐ Sticker to add to drug chart and/or medical notes
- ☐ Page to add to medical notes
- ☐ Pocket-sized card
- ☐ Poster for e.g. treatment room
- ☐ Other

17

Is the current design and format of the IVOS criteria clear? (draft version emailed as PDF and available here as image) *

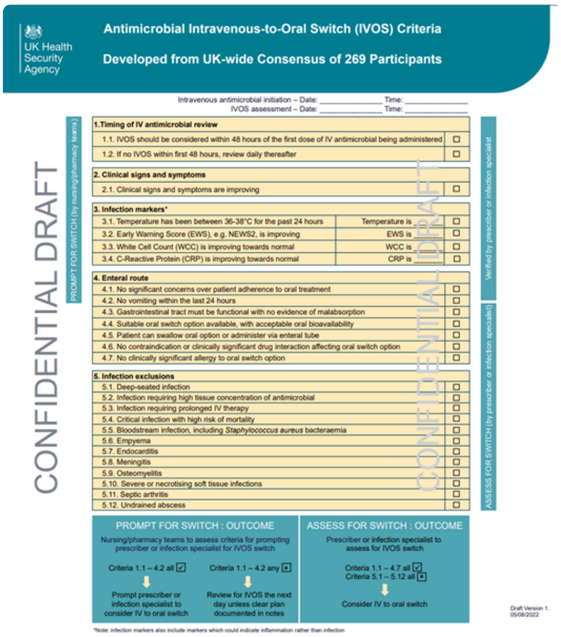

UK Health Security Agency

**Antimicrobial Intravenous-to-Oral Switch (IVOS) Criteria**

**Developed from UK-wide Consensus of 269 Participants**

CONFIDENTIAL DRAFT

Verified by prescriber or infection specialist

ASSESS FOR SWITCH (by prescriber or infection specialist)

**PROMPT FOR SWITCH : OUTCOME**

Nursing/pharmacy teams to assess criteria for prompting prescriber or infection specialist for IVOS switch

Criteria 1.1 – 4.2 all ☑ → Prompt prescriber or infection specialist to consider IV to oral switch

Criteria 1.1 – 4.2 any ☒ → Review for IVOS the next day unless clear plan documented in notes

**ASSESS FOR SWITCH : OUTCOME**

Prescriber or infection specialist to assess for IVOS switch

Criteria 1.1 – 4.7 all ☑

Criteria 5.1 – 5.12 all ☒

→ Consider IV to oral switch

Draft Version 1.
05/08/2022

*Note: infection markers also include markers which could indicate inflammation rather than infection

- ◯ Yes
- ◯ Somewhat
- ◯ No

18

Please use the box below for any feedback on the draft version of the IVOS tool, including suggestions for improvement.

#### Equality and diversity monitoring

19

Please indicate your gender.
*If 'Other', please self describe your gender.* *

- ◯ Female
- ◯ Male
- ◯ Prefer not to say
- ◯ Other

20

Please select what best describes your ethnic origin.
*If 'Other', please self describe your ethnic origin.* *

- ◯ White (including British, Irish, any other White background)
- ◯ Asian or Asian British (Indian, Pakistani, any other Asian background)
- ◯ Black or Black British (Caribbean, African, any other Black background)
- ◯ Mixed (White & Asian, White & Black, any other mixed background)
- ◯ Other ethnic groups (Chinese, any other ethnic group)
- ◯ Prefer not to say
- ◯ Other

#### Final comments

21

Thank you for your time in completing this survey. Please use the box below for any final comments.

22

Have you participated in any of the previous Steps 1-3 of the Delphi consensus-gathering process? *

- ◯ Yes
- ◯ No
- ◯ I can't remember

23

Are you able to attend the Delphi Step 4) Workshop scheduled for Wednesday 17 August 2022 from 11:00-12:30hrs?

- ◯ Yes
- ◯ No
- ◯ Maybe

24

Would you like to be contacted with the results of this study and involved in other Antimicrobial Stewardship projects? *

- ◯ Yes
- ◯ No

25

Please provide your email address (this information will be disaggregated and the rest of your survey responses will remain anonymous). *

This content is neither created nor endorsed by Microsoft. The data you submit will be sent to the form owner.

Microsoft Forms
