## Supplementary material for "A Delphi Process for Reaching Nationwide Consensus on Antimicrobial Intravenous-to-oral Switch Criteria and Development of an Antimicrobial Intravenous-to-oral Switch Decision Aid": Delphi Step One_Pilot1st Round_Questionnaire

### PILOT/1st ROUND: Questionnaire on Developing an Antimicrobial IV Switch Tool for Adults

Thank you for participating in this questionnaire as part of a consensus gathering (Delphi) process to inform an evidence-based, national antimicrobial intravenous (IV) switch tool for adults. The information you provide will remain anonymous.

The 'Start Smart – Then Focus' antimicrobial stewardship toolkit outlines five antimicrobial prescribing decision options, one of which is to switch antibiotics from intravenous to oral therapy in line with local policies (PHE, 2015). The literature outlines numerous benefits for IV switch to oral; including decreased risk of catheter-related infections, reduced costs and increased patient mobility and comfort (Nguyen *et al.*, 2021). In clinically stable patients, studies report that a timely IV switch is safe and of equal efficacy to the full course of IV therapy (McCarthy and Avent, 2020), with no negative impact on patient outcome (Wongkamhla *et al.*, 2020).

This questionnaire includes antimicrobial IV switch criteria from 45 local Trust or Health Board policies across the UK, the literature and expert opinion. In collaboration with Dr Kieran Hand (National Pharmacy and Prescribing Clinical Lead, NHSEI), we invite you to appraise the criteria in terms of their clinical relevance for safe and effective IV switch and their ease of assessment in the clinical setting of hospitalised adult patients.

The form will take approximately 15 minutes to complete and will be open until **Thursday 10th March 2022**. **(Extended to 15th March 2022)**

Please direct any queries to:

Eleanor Harvey – Chief Pharmaceutical Officer Clinical Fellow, UKHSA
Dr Diane Ashiru-Oredope – Chair, English Surveillance Programme for Antimicrobial Utilisation and Resistance and UKHSA's Lead Pharmacist for HCAI, AMR, AMU, Fungal and Sepsis Division

#### Consent

Consent form for professionals in the Delphi process to inform an evidence-based, national antimicrobial IV switch tool for adults. The information you provide will remain anonymous.

1. I consent to participating in this questionnaire and understand that any information I provide will be used to help inform an evidence-based, national antimicrobial IV switch tool for adults *

◯ Yes

◯ No

### About you

Where AM = Antimicrobial; ID = Infectious Diseases

2. Please provide your name *

3. Please provide your email address *

4. What is your profession? *

- ◯ Microbiologist/ID Physician
- ◯ General Physician
- ◯ Surgeon
- ◯ Dentist or Dental Nurse
- ◯ AM/ID Nurse
- ◯ General Nurse/Midwife
- ◯ Specialist AM/ID Pharmacist
- ◯ General Pharmacist
- ◯ Allied Health Professional
- ◯ AM/ID Clinical Researcher
- ◯ General Clinical Researcher
- ◯ Other

5. How many years have you been practising within your profession? *

6. If an AM/ID specialist, for how long have you been in your specialist role? *

- 0-1 year
- 1-2 years
- 2-5 years
- 5-10 years
- 10+ years
- Not applicable

7. Which of the following describes your main area(s) of work? *

- [ ] NHS Acute Trust
- [ ] Health Board
- [ ] NHSEI
- [ ] UKHSA
- [ ] Other Arms Length Body/Organisation
- [ ] Mental Health
- [ ] Community
- [ ] Academic
- [ ] Government
- [ ] General Practice/CCG
- [ ] Clinical Fellow (national/regional)
- [ ] Other

8. If you are a Clinical Fellow, please state your role and organisation prior to the fellowship (e.g. clinical pharmacist at an acute hospital)

9. Which country do you work in? *

- England
- Northern Ireland
- Scotland
- Wales
- Other

10. Which region do you work in? *

- East of England
- London
- Midlands
- North East and Yorkshire
- North West
- South East
- South West
- National
- Not applicable

11. What is your gender? *

- Male
- Female
- Prefer not to say

12. PILOT/1st ROUND: Please provide comments on the content of this page, including the question number

### Antimicrobial IV switch criteria

Respondents are asked to appraise each criterion via a 5-point Likert scale, with additional options of 'Not applicable' or 'I don't know'.

The questionnaire presents IV switch criteria in 5 sections:

- Timing of IV antimicrobial review
- Clinical signs and symptoms
- Infection markers
- Enteral route
- Infection exceptions

Each section is presented twice:
1) Please consider the clinical **relevance** of criteria for safe and effective IV switch. For each criterion: is it clinically significant and does it need to be met for a safe and effective IV switch? does it apply to hospitalised adults on IV antimicrobials?
2) Please consider the **ease** with which criteria can be assessed in the clinical setting. For each criterion: how achievable is it in practice?

[illegible]

13. For clinically stable patients, in your experience what impact has antimicrobial IV switch had on patient outcomes? *

- ○ Positive
- ○ Negative
- ○ Neither positive nor negative
- ○ I don't know

14. Timing of IV antimicrobial review: How clinically relevant is it to review IV antimicrobials within each specified timeframe? *

| | Highly relevant | Relevant | More or less relevant | Not relevant | Not at all relevant | Not applical |
|---|---|---|---|---|---|---|
| a. Review antimicrobial within **24 hours** | ○ | ○ | ○ | ○ | ○ | ○ |
| b. Review antimicrobial within **24-48 hours** | ○ | ○ | ○ | ○ | ○ | ○ |
| c. Review antimicrobial within **48 hours** | ○ | ○ | ○ | ○ | ○ | ○ |
| d. Review antimicrobial within **48-72 hours** | ○ | ○ | ○ | ○ | ○ | ○ |

15. Timing of IV antimicrobial review: How easy is it for the clinical team to review IV antimicrobials within each specified timeframe? *

| | Very easy | Easy | More or less easy | Not easy | Not at all easy | Not applical |
|---|---|---|---|---|---|---|
| a. Review antimicrobial within **24 hours** | ◯ | ◯ | ◯ | ◯ | ◯ | ◯ |
| b. Review antimicrobial within **24-48 hours** | ◯ | ◯ | ◯ | ◯ | ◯ | ◯ |
| c. Review antimicrobial within **48 hours** | ◯ | ◯ | ◯ | ◯ | ◯ | ◯ |
| d. Review antimicrobial within **48-72 hours** | ◯ | ◯ | ◯ | ◯ | ◯ | ◯ |

16. Clinical signs and symptoms: How clinically relevant is this criterion for safe and effective IV switch? *

| | Highly relevant | Relevant | More or less relevant | Not relevant | Not at all relevant | Not applical |
|---|---|---|---|---|---|---|
| a. Clinical signs and symptoms should be improving | ◯ | ◯ | ◯ | ◯ | ◯ | ◯ |

17. Clinical signs and symptoms: How easy is it for the clinical team to assess this criterion for IV switch? *

| | Very easy | Easy | More or less easy | Not easy | Not at all easy | Not applical |
|---|---|---|---|---|---|---|
| a. Clinical signs and symptoms should be improving | ◯ | ◯ | ◯ | ◯ | ◯ | ◯ |

18. Infection markers: How clinically relevant is each criterion for safe and effective IV switch? *

| | Highly relevant | Relevant | More or less relevant | Not relevant | Not at all relevant | Not applical |
|---|---|---|---|---|---|---|
| a. Temperature should be between 36-38°C | ◯ | ◯ | ◯ | ◯ | ◯ | ◯ |
| b. Temperature should be between 36-38°C for past **24 hours** | ◯ | ◯ | ◯ | ◯ | ◯ | ◯ |
| c. Heart rate should be below 90 beats per minute | ◯ | ◯ | ◯ | ◯ | ◯ | ◯ |
| d. Heart rate should be below 90 beats per minute for past **12 hours** | ◯ | ◯ | ◯ | ◯ | ◯ | ◯ |
| | Highly relevant | Relevant | More or less relevant | Not relevant | Not at all relevant | Not applical |

| | Highly relevant | Relevant | More or less relevant | Not relevant | Not at all relevant | Not applical |
|---|---|---|---|---|---|---|
| e. Heart rate should be below 90 beats per minute for past **24 hours** | ○ | ○ | ○ | ○ | ○ | ○ |
| f. Blood pressure should be stable | ○ | ○ | ○ | ○ | ○ | ○ |
| g. Blood pressure should be stable for past **24 hours** | ○ | ○ | ○ | ○ | ○ | ○ |
| h. Respiratory rate should be below 20 breaths per minute | ○ | ○ | ○ | ○ | ○ | ○ |
| i. Respiratory rate should be below 20 breaths per minute for past **24 hours** | ○ | ○ | ○ | ○ | ○ | ○ |
| j. White cell count should be normalising | ○ | ○ | ○ | ○ | ○ | ○ |
| k. White cell count should be between 4 and 12 x10^9/L | ○ | ○ | ○ | ○ | ○ | ○ |
| l. White cell count should be between 4 and 12 x10^9/L or normalising | ○ | ○ | ○ | ○ | ○ | ○ |
| m. C-reactive | | | | | | |

| | | | | | | |
|---|---|---|---|---|---|---|
| protein should be normalising | ○ | ○ | ○ | ○ | ○ | ○ |
| n. C-reactive protein does not reflect severity of illness or the need for IV antibiotics, and may remain elevated as the infection improves | ○ | ○ | ○ | ○ | ○ | ○ |

19. Infection markers: How easy is it for the clinical team to assess each criterion for IV switch? *

| | Very easy | Easy | More or less easy | Not easy | Not at all easy | Not applical |
|---|---|---|---|---|---|---|
| a. Temperature should be between 36-38°C | ○ | ○ | ○ | ○ | ○ | ○ |
| b. Temperature should be between 36-38°C for past **24 hours** | ○ | ○ | ○ | ○ | ○ | ○ |
| c. Heart rate should be below 90 beats per minute | ○ | ○ | ○ | ○ | ○ | ○ |
| d. Heart rate should be below 90 beats per minute for past **12 hours** | ○ | ○ | ○ | ○ | ○ | ○ |
| | Very easy | Easy | More or less easy | Not easy | Not at all easy | Not applical |

| | Very easy | Easy | More or less easy | Not easy | Not at all easy | Not applical |
|---|---|---|---|---|---|---|
| e. Heart rate should be below 90 beats per minute for past **24 hours** | ○ | ○ | ○ | ○ | ○ | ○ |
| f. Blood pressure should be stable | ○ | ○ | ○ | ○ | ○ | ○ |
| g. Blood pressure should be stable for past **24 hours** | ○ | ○ | ○ | ○ | ○ | ○ |
| h. Respiratory rate should be below 20 breaths per minute | ○ | ○ | ○ | ○ | ○ | ○ |
| i. Respiratory rate should be below 20 breaths per minute for past **24 hours** | ○ | ○ | ○ | ○ | ○ | ○ |
| j. White cell count should be normalising | ○ | ○ | ○ | ○ | ○ | ○ |
| k. White cell count should be between 4 and 12 x10^9/L | ○ | ○ | ○ | ○ | ○ | ○ |
| l. White cell count should be between 4 and 12 x10^9/L or normalising | ○ | ○ | ○ | ○ | ○ | ○ |
| m. C-reactive [illegible] | | | | | | |

| |
|---|
| protein should be normalising |
| n. C-reactive protein does not reflect severity of illness or the need for IV antibiotics, and may remain elevated as the infection improves |

20. Enteral route: How clinically relevant is each criterion for safe and effective IV switch? *

| | Highly relevant | Relevant | More or less relevant | Not relevant | Not at all relevant | Not applical |
|---|---|---|---|---|---|---|
| a. Gastrointestinal tract must be functional | ○ | ○ | ○ | ○ | ○ | ○ |
| b. Patient can tolerate/ swallow oral option | ○ | ○ | ○ | ○ | ○ | ○ |
| c. No evidence of malabsorption | ○ | ○ | ○ | ○ | ○ | ○ |
| d. No vomiting | ○ | ○ | ○ | ○ | ○ | ○ |
| e. There should be a suitable oral option available | ○ | ○ | ○ | ○ | ○ | ○ |
| f. Check for drug interactions of oral option with patient's other medication | ○ | ○ | ○ | ○ | ○ | ○ |
| g. Check for allergies to oral option | ○ | ○ | ○ | ○ | ○ | ○ |
| h. Check patient adherence to oral option | ○ | ○ | ○ | ○ | ○ | ○ |

21. Enteral route: How easy is it for the clinical team to assess each criterion for IV switch? *

| | Very easy | Easy | More or less easy | Not easy | Not at all easy | Not applical |
|---|---|---|---|---|---|---|
| a. Gastrointestinal tract must be functional | ◯ | ◯ | ◯ | ◯ | ◯ | ◯ |
| b. Patient can tolerate/ swallow oral option | ◯ | ◯ | ◯ | ◯ | ◯ | ◯ |
| c. No evidence of malabsorption | ◯ | ◯ | ◯ | ◯ | ◯ | ◯ |
| d. No vomiting | ◯ | ◯ | ◯ | ◯ | ◯ | ◯ |
| e. There should be a suitable oral option available | ◯ | ◯ | ◯ | ◯ | ◯ | ◯ |
| f. Check for drug interactions of oral option with patient's other medication | ◯ | ◯ | ◯ | ◯ | ◯ | ◯ |
| g. Check for allergies to oral option | ◯ | ◯ | ◯ | ◯ | ◯ | ◯ |
| h. Check patient adherence to oral option | ◯ | ◯ | ◯ | ◯ | ◯ | ◯ |

22. Infection exceptions: How clinically relevant is each criterion to warrant exclusion from using this IV switch tool? (and for referral to clinician in charge of patient's care) *

| | Highly relevant | Relevant | More or less relevant | Not relevant | Not at all relevant | Not applical |
|---|---|---|---|---|---|---|
| a. Deep-seated infections | ○ | ○ | ○ | ○ | ○ | ○ |
| b. Infections requiring high tissue concentration | ○ | ○ | ○ | ○ | ○ | ○ |
| c. Infections requiring prolonged IV therapy | ○ | ○ | ○ | ○ | ○ | ○ |
| d. Critical infection with high risk of mortality | ○ | ○ | ○ | ○ | ○ | ○ |
| e. On microbiology advice | ○ | ○ | ○ | ○ | ○ | ○ |
| f. Endocarditis | ○ | ○ | ○ | ○ | ○ | ○ |
| g. Meningitis | ○ | ○ | ○ | ○ | ○ | ○ |
| h. Bacteraemia, including *Staph. aureus* | ○ | ○ | ○ | ○ | ○ | ○ |
| i. Immunocompromised | ○ | ○ | ○ | ○ | ○ | ○ |
| j. Abscess | ○ | ○ | ○ | ○ | ○ | ○ |

| | Highly relevant | Relevant | More or less relevant | Not relevant | Not at all relevant | Not applical |
|---|---|---|---|---|---|---|
| k. Severe or necrotising soft tissue | ○ | ○ | ○ | ○ | ○ | ○ |

| |
|---|
| infections |
| l. Infections of foreign bodies |
| m. Osteomyelitis |
| n. Septic arthritis |
| o. Empyema |

23. Infection exceptions: How easy is it for the clinical team to assess each criterion? *

| | Very easy | Easy | More or less easy | Not easy | Not at all easy | Not applical |
|---|---|---|---|---|---|---|
| a. Deep-seated infections | | | | | | |
| b. Infections requiring high tissue concentration | | | | | | |
| c. Infections requiring prolonged IV therapy | | | | | | |
| d. Critical infection with high risk of mortality | | | | | | |
| e. On microbiology advice | | | | | | |
| f. Endocarditis | | | | | | |
| g. Meningitis | Very easy | Easy | More or less easy | Not easy | Not at all easy | Not applical |

| | | | | | | |
|---|---|---|---|---|---|---|
| h. Bacteraemia, including *Staph. aureus* | ○ | ○ | ○ | ○ | ○ | ○ |
| i. Immunocompromised | ○ | ○ | ○ | ○ | ○ | ○ |
| j. Abscess | ○ | ○ | ○ | ○ | ○ | ○ |
| k. Severe or necrotising soft tissue infections | ○ | ○ | ○ | ○ | ○ | ○ |
| l. Infections of foreign bodies | ○ | ○ | ○ | ○ | ○ | ○ |
| m. Osteomyelitis | ○ | ○ | ○ | ○ | ○ | ○ |
| n. Septic arthritis | ○ | ○ | ○ | ○ | ○ | ○ |
| o. Empyema | ○ | ○ | ○ | ○ | ○ | ○ |

24. Please use the box below if you would like to suggest any criteria word amendments, please specify which criteria you are referring to (e.g. Question 14a)

25. Please use the box below if you would like to suggest any additional criteria that would add benefit to the national IV switch tool

26. For wider dissemination of questionnaire, which Likert scale wording is most appropriate to be able to determine whether criteria should be met for a safe and effective IV switch?

- ○ Clinically **relevant**
- ○ Clinically **significant**
- ○ Other

27. In your opinion, how feasible is it for the **nursing** team to prompt an IV antimicrobial review in relation to each of the 5 outlined sections? *

| | Very feasible | Feasible | More or less feasible | Not feasible | Not at all feasible | Not applical |
|---|---|---|---|---|---|---|
| Timing of IV antimicrobial review | ○ | ○ | ○ | ○ | ○ | ○ |
| Clinical signs and symptoms | ○ | ○ | ○ | ○ | ○ | ○ |
| Infection markers | ○ | ○ | ○ | ○ | ○ | ○ |
| Enteral route | ○ | ○ | ○ | ○ | ○ | ○ |
| Infection exceptions | ○ | ○ | ○ | ○ | ○ | ○ |

28. In your opinion, how feasible is it for the **pharmacy** team to prompt an IV antimicrobial review in relation to each of the 5 outlined sections? *

| | Very feasible | Feasible | More or less feasible | Not feasible | Not at all feasible | Not applical |
|---|---|---|---|---|---|---|
| Timing of IV antimicrobial review | ○ | ○ | ○ | ○ | ○ | ○ |
| Clinical signs and symptoms | ○ | ○ | ○ | ○ | ○ | ○ |
| Infection markers | ○ | ○ | ○ | ○ | ○ | ○ |
| Enteral route | ○ | ○ | ○ | ○ | ○ | ○ |
| Infection exceptions | ○ | ○ | ○ | ○ | ○ | ○ |

29. PILOT/1st ROUND: Please provide comments on the content of this page, including the question number

### Electronic Prescribing and Medicines Administration (ePMA)

30. In your clinical area of work, do you use ePMA systems? *

- Yes
- No
- Not applicable

31. Has ePMA facilitated, hindered or not affected antimicrobial IV switch? *

- Facilitated
- Hindered
- Not affected
- I don't know

32. PILOT/1ST ROUND: Please provide comments on the content of this page, including the question number

#### Further comments

33. Thank you for your time in completing this survey. Please use the box below for any further comments

34. Can we contact you further regarding this project? *

- ◯ Yes
- ◯ No

This content is neither created nor endorsed by Microsoft. The data you submit will be sent to the form owner.

Microsoft Forms
