## Supplementary material for "A Delphi Process for Reaching Nationwide Consensus on Antimicrobial Intravenous-to-oral Switch Criteria and Development of an Antimicrobial Intravenous-to-oral Switch Decision Aid": Delphi Step Three_2nd Round_Questionnaire

### UK-wide Consensus for an Antimicrobial Intravenous-to-oral Switch (IVOS) Tool for Adults

We invite **healthcare professionals** (especially those who are patient facing) to complete this questionnaire to inform an evidence-based antimicrobial intravenous-to-oral switch (IVOS) tool for hospitalised adult patients.

This version of the questionnaire has been developed following feedback from 30 multidisciplinary healthcare professionals. We are now looking for 140 multidisciplinary healthcare professionals across the UK to take part in this consensus gathering Delphi process. **Please cascade to colleagues**.

The form will take approximately 12 minutes to complete and will be open until **Tuesday 14th June 2022**. **(Extended to 17th June 2022)**
Please ensure you click submit at the end of the form, otherwise none of your responses will be recorded.
The information you provide will remain anonymous.

Please direct any queries to:
Eleanor Harvey – Chief Pharmaceutical Officer Clinical Fellow, UKHSA
Dr Diane Ashiru-Oredope – Chair, English Surveillance Programme for Antimicrobial Utilisation and Resistance and UKHSA's Lead Pharmacist for HCAI, AMR, AMU, Fungal and Sepsis Division

**About this study**

The 'Start Smart – Then Focus' antimicrobial stewardship toolkit outlines five antimicrobial prescribing decision options, one of which is IVOS (PHE, 2015). The literature outlines numerous benefits for IVOS; including decreased risk of catheter-related infections, reduced costs and increased patient mobility and comfort (Nguyen *et al.*, 2021). In clinically stable patients, studies report that a timely IVOS is safe and of equal efficacy to the full course of IV therapy (McCarthy and Avent, 2020), with no negative impact on patient outcome (Wongkamhla *et al.*, 2020).

This questionnaire includes antimicrobial IVOS criteria from 45 local Trust or Health Board policies across the UK, the literature and expert opinion. In collaboration with Dr Kieran Hand (National Pharmacy and Prescribing Clinical Lead, NHSEI), we invite you to appraise the criteria for safe and effective antimicrobial IVOS in the clinical setting of hospitalised adult patients.

#### Consent

Consent form for professionals in the Delphi process to inform an evidence-based antimicrobial intravenous-to-oral switch (IVOS) tool for hospitalised adult patients. The information you [illegible]

1. I consent to participating in this questionnaire and understand that any information I provide will be used to help inform an evidence-based antimicrobial IVOS tool for hospitalised adult patients. *

   ◯ Yes

   ◯ No

### About you

2. Are you actively involved in the clinical decision making, advising or implementing of antimicrobial IVOS? *

- ◯ Yes
- ◯ No
- ◯ Not currently

3. What is your profession?
*If 'Other', please include profession.* *

- ◯ Medical Microbiologist/Infection Diseases Doctor
- ◯ General Physician
- ◯ Surgeon
- ◯ Dentist or Dental Nurse
- ◯ Nurse (Antimicrobial Stewardship)
- ◯ Nurse (Infection Prevention Control)
- ◯ Nurse/Midwife (General or non-Infection Specialist)
- ◯ Pharmacist (Antimicrobial or Infection Specialist)
- ◯ Pharmacist (General or non-Infection Specialist)
- ◯ Allied Health Professional
- ◯ Clinical Researcher (Infection Specialist)
- ◯ Clinical Researcher (General)
- ◯ Healthcare Scientist
- ◯ Other

4. How many years have you been practising within your profession? *

5. If an infection specialist, for how long have you been in your specialist role? *

- ○ 0-1 year
- ○ 1-2 years
- ○ 2-5 years
- ○ 5-10 years
- ○ 10+ years
- ○ Not applicable

6. Which of the following describes your main area(s) of work? *

- [ ] NHS Non-teaching Acute Trust
- [ ] NHS Specialist Acute Trust
- [ ] NHS Teaching Acute Trust
- [ ] Independent Hospital
- [ ] Community Health Trust
- [ ] Mental Health Trust
- [ ] NHSEI
- [ ] UKHSA
- [ ] Other Arms Length Body/Organisation
- [ ] Academia
- [ ] Government
- [ ] General Practice/CCG
- [ ] Other

7. Which country do you work in? *

- ○ England
- ○ Northern Ireland
- ○ Scotland
- ○ Wales
- ○ Other

8. Which region do you work in? *

- East of England
- London
- Midlands
- North East and Yorkshire
- North West
- South East
- South West
- National

9. Please provide the first half of your organisation's post code (to ensure UK-wide representation).

#### Your experience

10. For clinically stable patients, what has been your experience of timely antimicrobial IVOS on patient outcomes? *

- ◯ Positive
- ◯ Negative
- ◯ Neither positive nor negative
- ◯ Both positive and negative
- ◯ I don't know
- ◯ Other

11. In the past 2 weeks, what IVOS barriers have you encountered? Please select all that apply.
*If you have had other barriers, please select 'Other' and provide a short explanation.*

- ☐ Lack of senior agreement to IVOS
- ☐ Lack of time to review patient for IVOS suitability
- ☐ Lack of decision support tool for IVOS (e.g. checklist)
- ☐ No suitable oral option available
- ☐ Culture results unknown

12. Please use the box below for any further comments regarding your experience.

### Antimicrobial IVOS criteria

The questionnaire presents IVOS criteria in 5 sections:

1) Timing of IV antimicrobial review
2) Clinical signs and symptoms
3) Infection markers
4) Enteral route
5) Infection exclusions

For each of the first 4 sections, assume the patient has an infection that is <u>not excluded</u> from IVOS.

#### 13. Timing of IV antimicrobial review: To what extent do you agree with the following timeframes of when IVOS should be considered? *

| | Strongly agree | Agree | Neither agree nor disagree | Disagree | Strongly disagree | Not applical |
|---|---|---|---|---|---|---|
| a. IVOS should be considered **any time after** the first dose of IV antimicrobial is administered | ○ | ○ | ○ | ○ | ○ | ○ |
| b. IVOS should be considered **within 24 hours** of the first dose of IV antimicrobial being administered | ○ | ○ | ○ | ○ | ○ | ○ |
| c. IVOS should be considered **24 hours after** the first dose of IV antimicrobial | ○ | ○ | ○ | ○ | ○ | ○ |

Strongly agree Agree Neither agree nor disagree Disagree Strongly disagree Not applical

| | | | | | | |
|---|---|---|---|---|---|---|
| is administered | | | | | | |
| d. IVOS should be considered **within 48 hours** of the first dose of IV antimicrobial being administered | ◯ | ◯ | ◯ | ◯ | ◯ | ◯ |
| e. IVOS should be considered **48 hours after** the first dose of IV antimicrobial is administered | ◯ | ◯ | ◯ | ◯ | ◯ | ◯ |

14. Timing of IV antimicrobial review: How often should IVOS be considered after initial review?
*If 'Other', please include proposed timing.* *

◯ Daily thereafter

◯ Every two days thereafter

◯ I don't know

◯ Other

15. Please use the box below for any further comments regarding timing of IV antimicrobial review.

16. Clinical signs and symptoms: To what extent do you agree with the following criteria as being essential for a safe and effective IVOS? *

| | Strongly agree | Agree | Neither agree nor disagree | Disagree | Strongly disagree | Not applical |
|---|---|---|---|---|---|---|
| a. Clinical signs and symptoms are **improving** | ○ | ○ | ○ | ○ | ○ | ○ |
| b. Clinical signs and symptoms are **not worsening** | ○ | ○ | ○ | ○ | ○ | ○ |

17. Infection markers: To what extent do you agree with the following criteria as being essential for a safe and effective IVOS? *

| | Strongly agree | Agree | Neither agree nor disagree | Disagree | Strongly disagree | Not applical |
|---|---|---|---|---|---|---|
| a. Temperature is between 36-38°C | ○ | ○ | ○ | ○ | ○ | ○ |
| b. Temperature is between 36-38°C for the past **24 hours** | ○ | ○ | ○ | ○ | ○ | ○ |
| c. Early Warning Score (e.g. MEWS, NEWS2) is **improving** | ○ | ○ | ○ | ○ | ○ | ○ |

Strongly agree | Agree | Neither agree nor disagree | Disagree | Strongly disagree | Not applical

d. Early [illegible]

| | | | | | | |
|---|---|---|---|---|---|---|
| d. Early Warning Score (e.g. MEWS, NEWS2) is **not worsening** | ○ | ○ | ○ | ○ | ○ | ○ |
| e. White Cell Count is **improving** | ○ | ○ | ○ | ○ | ○ | ○ |
| f. White Cell Count is **not worsening** | ○ | ○ | ○ | ○ | ○ | ○ |
| g. C-Reactive Protein is **improving** | ○ | ○ | ○ | ○ | ○ | ○ |
| h. C-Reactive Protein is **not worsening** | ○ | ○ | ○ | ○ | ○ | ○ |

18. Enteral route: To what extent do you agree with the following criteria as being essential for a safe and effective IVOS? *

| | Strongly agree | Agree | Neither agree nor disagree | Disagree | Strongly disagree | Not applical |
|---|---|---|---|---|---|---|
| a. Gastrointestin al tract must be functional | ○ | ○ | ○ | ○ | ○ | ○ |
| b. Patient can tolerate/ swallow oral option | ○ | ○ | ○ | ○ | ○ | ○ |
| c. No evidence of malabsorptio n | ○ | ○ | ○ | ○ | ○ | ○ |
| d. No vomiting within the | ○ | ○ | ○ | ○ | ○ | ○ |

Strongly agree Agree Neither agree nor disagree Disagree Strongly disagree Not applical

| | | | | | | |
|---|---|---|---|---|---|---|
| within the last 24 hours | | | | | | |
| e. Suitable oral switch option available | ○ | ○ | ○ | ○ | ○ | ○ |
| f. No contraindication or clinically significant drug interaction affecting oral switch option | ○ | ○ | ○ | ○ | ○ | ○ |
| g. No clinically significant allergy to oral switch option | ○ | ○ | ○ | ○ | ○ | ○ |
| h. No significant concerns over patient adherence to oral switch option | ○ | ○ | ○ | ○ | ○ | ○ |

19. Infection exclusions: To what extent do you agree that the following infections must be excluded from an early IVOS (e.g. within 48 hours) in the absence of specialist advice? *

| | Strongly agree | Agree | Neither agree nor disagree | Disagree | Strongly disagree | Not applical |
|---|---|---|---|---|---|---|
| a. Deep-seated infection | ○ | ○ | ○ | ○ | ○ | ○ |
| b. Infection requiring high tissue concentration | ○ | ○ | ○ | ○ | ○ | ○ |

Strongly agree Agree Neither agree nor disagree Disagree Strongly disagree Not applical

| | | | | | | |
|---|---|---|---|---|---|---|
| c. Infection requiring prolonged IV therapy | ◯ | ◯ | ◯ | ◯ | ◯ | ◯ |
| d. Critical infection with high risk of mortality | ◯ | ◯ | ◯ | ◯ | ◯ | ◯ |
| e. Bacteraemia, including Sta ph. aureus | ◯ | ◯ | ◯ | ◯ | ◯ | ◯ |
| f. Empyema | ◯ | ◯ | ◯ | ◯ | ◯ | ◯ |
| g. Endocarditis | ◯ | ◯ | ◯ | ◯ | ◯ | ◯ |
| h. Meningitis | ◯ | ◯ | ◯ | ◯ | ◯ | ◯ |
| i. Osteomyelitis | ◯ | ◯ | ◯ | ◯ | ◯ | ◯ |
| j. Septic arthritis | ◯ | ◯ | ◯ | ◯ | ◯ | ◯ |
| k. Severe or necrotising soft tissue infections | ◯ | ◯ | ◯ | ◯ | ◯ | ◯ |
| l. Undrained abscess | ◯ | ◯ | ◯ | ◯ | ◯ | ◯ |

20. Please use the box below if you would like to suggest any criteria word amendments. Please include both the original criteria and what your recommended changes are.

21. Please use the box below if you would like to suggest any additional criteria that would add benefit to the IVOS tool.

22. In your opinion, how feasible is it for the **nursing** team to prompt an IV antimicrobial review in relation to each of the 5 outlined sections? *

| | Very feasible | Feasible | Somewhat feasible | Not feasible | Not at all feasible | Not applical |
|---|---|---|---|---|---|---|
| Timing of IV antimicrobial review | ○ | ○ | ○ | ○ | ○ | ○ |
| Clinical signs and symptoms | ○ | ○ | ○ | ○ | ○ | ○ |
| Infection markers | ○ | ○ | ○ | ○ | ○ | ○ |
| Enteral route | ○ | ○ | ○ | ○ | ○ | ○ |
| Infection exclusions | ○ | ○ | ○ | ○ | ○ | ○ |

23. In your opinion, how feasible is it for the **pharmacy** team to prompt an IV antimicrobial review in relation to each of the 5 outlined sections? *

| | Very feasbile | Feasible | Somewhat feasible | Not feasible | Not at all feasible | Not applical |
|---|---|---|---|---|---|---|
| Timing of IV antimicrobial review | ◯ | ◯ | ◯ | ◯ | ◯ | ◯ |
| Clinical signs and symptoms | ◯ | ◯ | ◯ | ◯ | ◯ | ◯ |
| Infection markers | ◯ | ◯ | ◯ | ◯ | ◯ | ◯ |
| Enteral route | ◯ | ◯ | ◯ | ◯ | ◯ | ◯ |
| Infection exclusions | ◯ | ◯ | ◯ | ◯ | ◯ | ◯ |

24. Please use the box below for any further comments regarding feasibility of nursing/pharmacy team to prompt an IV antimicrobial review.

### Electronic Prescribing and Medicines Administration (ePMA)

25. In your clinical area of work, do you use ePMA systems? *

- ◯ Yes
- ◯ No
- ◯ Not applicable

26. Has ePMA facilitated, hindered or not affected antimicrobial IVOS? *

- ◯ Facilitated
- ◯ Hindered
- ◯ Not affected
- ◯ I don't know

27. Please use the box below for any further comments regarding ePMA and antimicrobial IVOS.

#### Equality and diversity monitoring

28. Please indicate your gender.
    *If 'Other', please self describe your gender.* *

- ◯ Female
- ◯ Male
- ◯ Prefer not to say
- ◯ Other

29. Please select what best describes your ethnic origin.
    *If 'Other', please self describe your ethnic origin.* *

- ◯ White (including British, Irish, any other White background)
- ◯ Asian or Asian British (Indian, Pakistani, any other Asian background)
- ◯ Black or Black British (Caribbean, African, any other Black background)
- ◯ Mixed (White & Asian, White & Black, any other mixed background)
- ◯ Other ethnic groups (Chinese, any other ethnic group)
- ◯ Prefer not to say
- ◯ Other

#### Final comments

30. Thank you for your time in completing this questionnaire. Please use the box below for any final comments.

31. Would you like to be contacted with the results of this study and involved in other Antimicrobial Stewardship projects? *

- ○ Yes
- ○ No

32. Please provide your email address (this information will be disaggregated and the rest of your questionnaire responses will remain anonymous). *

This content is neither created nor endorsed by Microsoft. The data you submit will be sent to the form owner.

Microsoft Forms
