## Supplementary material for "A Delphi Process for Reaching Nationwide Consensus on Antimicrobial Intravenous-to-oral Switch Criteria and Development of an Antimicrobial Intravenous-to-oral Switch Decision Aid": Delphi_AGREE Reporting Checklist

AGREE
REPORTING CHECKLIST

### AGREE Reporting Checklist

2016

*This checklist is intended to guide the reporting of clinical practice guidelines.*

| CHECKLIST ITEM AND DESCRIPTION | REPORTING CRITERIA | Page # |
|---|---|---|
| ***DOMAIN 1: SCOPE AND PURPOSE*** | | |
| **1. OBJECTIVES**<br>*Report the overall objective(s) of the guideline. The expected health benefits from the guideline are to be specific to the clinical problem or health topic.* | ☑ Health intent(s) (i.e., prevention, screening, diagnosis, treatment, etc.)<br>☑ Expected benefit(s) or outcome(s)<br>☑ Target(s) (e.g., patient population, society) | 3 |
| **2. QUESTIONS**<br>*Report the health question(s) covered by the guideline, particularly for the key recommendations.* | ☑ Target population<br>☑ Intervention(s) or exposure(s)<br>☐ Comparisons (if appropriate)<br>☑ Outcome(s)<br>☑ Health care setting or context | 4 |
| **3. POPULATION**<br>*Describe the population (i.e., patients, public, etc.) to whom the guideline is meant to apply.* | ☑ Target population, sex and age<br>☑ Clinical condition (if relevant)<br>☐ Severity/stage of disease (if relevant)<br>☐ Comorbidities (if relevant)<br>☑ Excluded populations (if relevant) | 3, 18 |
| ***DOMAIN 2: STAKEHOLDER INVOLVEMENT*** | | |
| **4. GROUP MEMBERSHIP**<br>*Report all individuals who were involved in the development process. This may include members of the steering group, the research team involved in selecting and reviewing/rating the evidence and individuals involved in formulating the final recommendations.* | ☐ Name of participant<br>☑ Discipline/content expertise (e.g., neurosurgeon, methodologist)<br>☑ Institution (e.g., St. Peter's hospital)<br>☑ Geographical location (e.g., Seattle, WA)<br>☑ A description of the member's role in the guideline development group | 8-14 |
| **5. TARGET POPULATION PREFERENCES AND VIEWS**<br>*Report how the views and preferences of the target population were sought/considered and what the resulting outcomes were.* | ☑ Statement of type of strategy used to capture patients'/publics' views and preferences (e.g., participation in the guideline development group, literature review of values and preferences)<br>☑ Methods by which preferences and views were sought (e.g., evidence from literature, surveys, focus groups)<br>☑ Outcomes/information gathered on patient/public information<br>☑ How the information gathered was used to inform the guideline development process and/or formation of the recommendations | 4-8 |
| **6. TARGET USERS**<br>*Report the target (or intended) users of the guideline.* | ☑ The intended guideline audience (e.g. specialists, family physicians, patients, clinical or institutional leaders/administrators)<br>☑ How the guideline may be used by its target audience (e.g., to inform clinical decisions, to inform policy, to inform standards of care) | 3-4 |

| DOMAIN 3: RIGOUR OF DEVELOPMENT | | |
|---|---|---|
| **7. SEARCH METHODS**<br>*Report details of the strategy used to search for evidence.* | ☐ Named electronic database(s) or evidence source(s) where the search was performed (e.g., MEDLINE, EMBASE, PsychINFO, CINAHL)<br>☐ Time periods searched (e.g., January 1, 2004 to March 31, 2008)<br>☐ Search terms used (e.g., text words, indexing terms, subheadings)<br>☐ Full search strategy included (e.g., possibly located in appendix) | 4 |
| **8. EVIDENCE SELECTION CRITERIA**<br>*Report the criteria used to select (i.e., include and exclude) the evidence. Provide rationale, where appropriate.* | ☑ Target population (patient, public, etc.) characteristics<br>☑ Study design<br>☐ Comparisons (if relevant)<br>☑ Outcomes<br>☐ Language (if relevant)<br>☑ Context (if relevant) | 4-5 |
| **9. STRENGTHS & LIMITATIONS OF THE EVIDENCE**<br>*Describe the strengths and limitations of the evidence. Consider from the perspective of the individual studies and the body of evidence aggregated across all the studies. Tools exist that can facilitate the reporting of this concept.* | ☐ Study design(s) included in body of evidence<br>☐ Study methodology limitations (sampling, blinding, allocation concealment, analytical methods)<br>☐ Appropriateness/relevance of primary and secondary outcomes considered<br>☐ Consistency of results across studies<br>☐ Direction of results across studies<br>☐ Magnitude of benefit versus magnitude of harm<br>☐ Applicability to practice context | NA |
| **10. FORMULATION OF RECOMMENDATIONS**<br>*Describe the methods used to formulate the recommendations and how final decisions were reached. Specify any areas of disagreement and the methods used to resolve them.* | ☑ Recommendation development process (e.g., steps used in modified Delphi technique, voting procedures that were considered)<br>☑ Outcomes of the recommendation development process (e.g., extent to which consensus was reached using modified Delphi technique, outcome of voting procedures)<br>☑ How the process influenced the recommendations (e.g., results of Delphi technique influence final recommendation, alignment with recommendations and the final vote) | 8-16 |
| **11. CONSIDERATION OF BENEFITS AND HARMS**<br>*Report the health benefits, side effects, and risks that were considered when formulating the recommendations.* | ☑ Supporting data and report of benefits<br>☑ Supporting data and report of harms/side effects/risks<br>☑ Reporting of the balance/trade-off between benefits and harms/side effects/risks<br>☑ Recommendations reflect considerations of both benefits and harms/side effects/risks | 3, 16-17 |
| **12. LINK BETWEEN RECOMMENDATIONS AND EVIDENCE**<br>*Describe the explicit link between the recommendations and the evidence on which they are based.* | ☑ How the guideline development group linked and used the evidence to inform recommendations<br>☑ Link between each recommendation and key evidence (text description and/or reference list)<br>☑ Link between recommendations and evidence summaries and/or evidence tables in the results section of the guideline | Supp info |

| | | |
|---|---|---|
| **13. EXTERNAL REVIEW**<br>*Report the methodology used to conduct the external review.* | ☑ Purpose and intent of the external review (e.g., to improve quality, gather feedback on draft recommendations, assess applicability and feasibility, disseminate evidence)<br>☑ Methods taken to undertake the external review (e.g., rating scale, open-ended questions)<br>☑ Description of the external reviewers (e.g., number, type of reviewers, affiliations)<br>☑ Outcomes/information gathered from the external review (e.g., summary of key findings)<br>☑ How the information gathered was used to inform the guideline development process and/or formation of the recommendations (e.g., guideline panel considered results of review in forming final recommendations) | 8-16 |
| **14. UPDATING PROCEDURE**<br>*Describe the procedure for updating the guideline.* | ☑ A statement that the guideline will be updated<br>☑ Explicit time interval or explicit criteria to guide decisions about when an update will occur<br>☑ Methodology for the updating procedure | 18 |
| ***DOMAIN 4: CLARITY OF PRESENTATION*** | | |
| **15. SPECIFIC AND UNAMBIGUOUS RECOMMENDATIONS**<br>*Describe which options are appropriate in which situations and in which population groups, as informed by the body of evidence.* | ☑ A statement of the recommended action<br>☑ Intent or purpose of the recommended action (e.g., to improve quality of life, to decrease side effects)<br>☑ Relevant population (e.g., patients, public)<br>☐ Caveats or qualifying statements, if relevant (e.g., patients or conditions for whom the recommendations would not apply)<br>☐ If there is uncertainty about the best care option(s), the uncertainty should be stated in the guideline | 16 |
| **16. MANAGEMENT OPTIONS**<br>*Describe the different options for managing the condition or health issue.* | ☐ Description of management options<br>☐ Population or clinical situation most appropriate to each option | NA |
| **17. IDENTIFIABLE KEY RECOMMENDATIONS**<br>*Present the key recommendations so that they are easy to identify.* | ☑ Recommendations in a summarized box, typed in bold, underlined, or presented as flow charts or algorithms<br>☑ Specific recommendations grouped together in one section | 15-16, supp info |
| ***DOMAIN 5: APPLICABILITY*** | | |
| **18. FACILITATORS AND BARRIERS TO APPLICATION**<br>*Describe the facilitators and barriers to the guideline's application.* | ☑ Types of facilitators and barriers that were considered<br>☑ Methods by which information regarding the facilitators and barriers to implementing recommendations were sought (e.g., feedback from key stakeholders, pilot testing of guidelines before widespread implementation)<br>☑ Information/description of the types of facilitators and barriers that emerged from the inquiry (e.g., practitioners have the skills to deliver the recommended care, sufficient equipment is not available to ensure all eligible members of the | 12, 17 |

| | | |
|---|---|---|
| | population receive mammography)<br>☑ How the information influenced the guideline development process and/or formation of the recommendations | 12, 17 |
| **19. IMPLEMENTATION ADVICE/TOOLS**<br>*Provide advice and/or tools on how the recommendations can be applied in practice.* | ☑ Additional materials to support the implementation of the guideline in practice. For example:<br>☐ Guideline summary documents<br>☐ Links to check lists, algorithms<br>☐ Links to how-to manuals<br>☐ Solutions linked to barrier analysis (see Item 18)<br>▣ Tools to capitalize on guideline facilitators (see Item 18)<br>☐ Outcome of pilot test and lessons learned | Supp info |
| **20. RESOURCE IMPLICATIONS**<br>*Describe any potential resource implications of applying the recommendations.* | ☑ Types of cost information that were considered (e.g., economic evaluations, drug acquisition costs)<br>☑ Methods by which the cost information was sought (e.g., a health economist was part of the guideline development panel, use of health technology assessments for specific drugs, etc.)<br>☐ Information/description of the cost information that emerged from the inquiry (e.g., specific drug acquisition costs per treatment course)<br>☐ How the information gathered was used to inform the guideline development process and/or formation of the recommendations | 3, 18 |
| **21. MONITORING/ AUDITING CRITERIA**<br>*Provide monitoring and/or auditing criteria to measure the application of guideline recommendations.* | ☐ Criteria to assess guideline implementation or adherence to recommendations<br>☐ Criteria for assessing impact of implementing the recommendations<br>☐ Advice on the frequency and interval of measurement<br>☑ Operational definitions of how the criteria should be measured | 17 |
| ***DOMAIN 6: EDITORIAL INDEPENDENCE*** | | |
| **22. FUNDING BODY**<br>*Report the funding body's influence on the content of the guideline.* | ☑ The name of the funding body or source of funding (or explicit statement of no funding)<br>☑ A statement that the funding body did not influence the content of the guideline | 19 |
| **23. COMPETING INTERESTS**<br>*Provide an explicit statement that all group members have declared whether they have any competing interests.* | ☑ Types of competing interests considered<br>☑ Methods by which potential competing interests were sought<br>☑ A description of the competing interests<br>☑ How the competing interests influenced the guideline process and development of recommendations | 19 |

From:
Brouwers MC, Kerkvliet K, Spithoff K, on behalf of the AGREE Next Steps Consortium. The AGREE Reporting Checklist: a tool to improve reporting of clinical practice guidelines. *BMJ* 2016;352:i1152. doi: 10.1136/bmj.i1152.

For more information about the AGREE Reporting Checklist, please visit the AGREE Enterprise website at www.agreetrust.org.
