## Supplementary material for "A Delphi Process for Reaching Nationwide Consensus on Antimicrobial Intravenous-to-oral Switch Criteria and Development of an Antimicrobial Intravenous-to-oral Switch Decision Aid": Delphi_All Steps_Criteria Outcomes

**Table SX**. Criteria outcomes of each step in the Delphi process: Step One) Pilot/1^st^ round questionnaire, Step Two) Virtual meeting, Step Three) 2^nd^ round questionnaire and Step 4) Workshop.

| **IVOS criteria** | **Step One)**  **Pilot/1^st^ round questionnaire** | | | | | **Step Two) Virtual meeting** | **Step Three)**  **2^nd^ round questionnaire** | | | **Step Four) Workshop** |
| --- | --- | --- | --- | --- | --- | --- | --- | --- | --- | --- |
|  | ‘Relevance’ median | Percentage agreement (relevant and very relevant) (%) | ‘Ease’ median | Percentage agreement (easy and very easy) (%) | Outcome | Outcome | Median | Percentage agreement (agree and strongly agree) (%) | Outcome | Outcome |
| 1. **Timing of IV antimicrobial review** | | | | | | | | | | |
| a. Review antimicrobial within **24 hours** | 4 | 70.83 | 4 | 54.17 | Accepted | Rephrased into 1e |  |  |  |  |
| b. Review antimicrobial within **24-48 hours** | 4.5 | 91.67 | 4 | 66.67 | Accepted | Rephrased into 1g |  |  |  |  |
| c. Review antimicrobial within **48 hours** | 5 | 91.67 | 4 | 66.67 | Accepted | Rephrased into 1h |  |  |  |  |
| d. Review antimicrobial within **48-72 hours** | 5 | 87.50 | 4.5 | 75.00 | Accepted | Rephrased into 1i |  |  |  |  |
| e. IVOS should be considered **any time after** the first dose of IV antimicrobial is administered |  |  |  |  |  | New criteria proposed | 4 | 65.29 | Rejected |  |
| f. IVOS should be considered **within 24 hours** of the first dose of IV antimicrobial being administered |  |  |  |  |  | Result from rephrase 1a | 4 | 61.98 | Rejected |  |
| g. IVOS should be considered **24 hours after** the first dose of IV antimicrobial |  |  |  |  |  | Result from rephrase 1b | 4 | 69.83 | Rejected |  |
| h. IVOS should be considered **within 48 hours** of the first dose of IV antimicrobial being administered |  |  |  |  |  | Result from rephrase 1c | 4 | 78.10 | Accepted | Accepted |
| i. IVOS should be considered **48 hours after** the first dose of IV antimicrobial is administered |  |  |  |  |  | Result from rephrase 1d | 4 | 71.49 | Rejected |  |
| j. If no IVOS within first 48 hours, review daily thereafter |  |  |  |  |  | New criteria proposed | - | 88.02 | Accepted | Accepted |
| j. If no IVOS within first 48 hours, review every two days thereafter |  |  |  |  |  | New criteria proposed | - | 6.20 | Rejected |  |
| 2. **Clinical signs and symptoms** | | | | | | | | | | |
| a. Clinical signs and symptoms should be improving | 5 | 91.67 | 4 | 58.33 | Accepted | Rephrased into 2b |  |  |  |  |
| b. Clinical signs and symptoms are improving |  |  |  |  |  | Result from rephrase 2a | 5 | 95.04 | Accepted | Rephrased into 2d |
| c. Clinical signs and symptoms are not worsening |  |  |  |  |  | New criteria proposed | 3 | 36.78 | Rejected |  |
| d. Clinical signs and symptoms of infection are improving |  |  |  |  |  |  |  |  |  | Result from rephrase 3b |
| 3. **Infection markers** | | | | | | | | | | |
| a. Temperature should be between 36-38°C | 4 | 54.17 | 4.5 | 75.00 | Uncertain | Rephrased into 3o |  |  |  |  |
| b. Temperature should be between 36-38°C for past **24 hours** | 4 | 70.83 | 4.5 | 70.83 | Accepted | Rephrased into 3o |  |  |  |  |
| c. Heart rate should be below 90 beats per minute | 3 | 45.83 | 4.5 | 70.83 | Rejected | Rephrased into 3q-r |  |  |  |  |
| d. Heart rate should be below 90 beats per minute for past **12 hours** | 3 | 45.83 | 4 | 62.50 | Rejected | Rejected |  |  |  |  |
| e. Heart rate should be below 90 beats per minute for past **24 hours** | 3 | 41.67 | 4.5 | 66.67 | Rejected | Rejected |  |  |  |  |
| f. Blood pressure should be stable | 4 | 58.33 | 4 | 70.83 | Uncertain | Rephrased into 3q-r |  |  |  |  |
| g. Blood pressure should be stable for past **24 hours** | 4 | 58.33 | 4 | 66.67 | Uncertain | Rejected |  |  |  |  |
| h. Respiratory rate should be below 20 breaths per minute | 3.4 | 50.00 | 4 | 66.67 | Rejected | Rephrased into 3q-r |  |  |  |  |
| i. Respiratory rate should be below 20 breaths per minute for past **24 hours** | 4 | 54.17 | 4 | 62.50 | Uncertain | Rejected |  |  |  |  |
| j. White cell count should be normalising | 4 | 54.17 | 4 | 66.67 | Uncertain | Rephrased into s-t |  |  |  |  |
| k. White cell count should be between 4 and 12 x10^9/L | 3 | 29.17 | 4 | 62.50 | Rejected | Rejected |  |  |  |  |
| l. White cell count should be between 4 and 12 x10^9/L or normalising | 3 | 45.83 | 4 | 62.50 | Rejected | Rejected |  |  |  |  |
| m. C-reactive protein should be normalising | 3 | 29.17 | 4 | 66.67 | Rejected | Rephrased into 3u-v |  |  |  |  |
| n. C-reactive protein does not reflect severity of illness or the need for IV antibiotics, and may remain elevated as the infection improves | 3 | 45.83 | 3.5 | 50.00 | Rejected | Rejected |  |  |  |  |
| o. Temperature is between 36-38°C |  |  |  |  |  | Result from rephrase 3a | 3 | 47.52 | Rejected |  |
| p. Temperature is between 36-38°C for the past **24 hours** |  |  |  |  |  | Result from rephrase 3b | 4 | 75.21 | Accepted | Accepted |
| q. Early Warning Score (e.g. MEWS, NEWS2) is **improving** |  |  |  |  |  | Result from rephrase 3c-i | 4 | 87.19 | Accepted | Rephrased into 3w |
| r. Early Warning Score (e.g. MEWS, NEWS2) is **not worsening** |  |  |  |  |  | Result from rephrase 3c-i | 3 | 35.54 | Rejected |  |
| s. White Cell Count is **improving** |  |  |  |  |  | Result from rephrase 3j | 4 | 76.03 | Accepted | Rephrased into 3x |
| t. White Cell Count is **not worsening** |  |  |  |  |  | Result from rephrase 3j | 3 | 34.30 | Rejected |  |
| u. C-Reactive Protein is **improving** |  |  |  |  |  | Result from rephrase 3m | 4 | 73.55 | Accepted | Rephrased into 3y |
| v. C-Reactive Protein is **not worsening** |  |  |  |  |  | Result from rephrase 3m | 3 | 33.47 | Rejected |  |
| w. Early Warning Score is **decreasing** |  |  |  |  |  |  |  |  |  | Result from rephrase 3q |
| x. White Cell Count is **trending towards the normal range** |  |  |  |  |  |  |  |  |  | Result from rephrase 3s |
| y. C-Reactive Protein is **decreasing** |  |  |  |  |  |  |  |  |  | Result from rephrase 3u |
| 4. **Enteral route** | | | | | | | | | | |
| a. Gastrointestinal tract must be functional | 5 | 87.50 | 3 | 29.17 | Accepted | Accepted | 5 | 96.69 | Accepted | Rephrased into 4n |
| b. Patient can tolerate/ swallow oral option | 5 | 87.50 | 4 | 62.50 | Accepted | Accepted | 5 | 96.69 | Accepted | Rephrased into 4o |
| c. No evidence of malabsorption | 4 | 91.67 | 3 | 29.17 | Accepted | Accepted | 5 | 95.04 | Accepted | Rephrased into 4n |
| d. No vomiting | 4.5 | 91.67 | 4 | 75.00 | Accepted | Rephrased into 4i |  |  |  |  |
| e. There should be a suitable oral option available | 5 | 91.67 | 4 | 54.17 | Accepted | Rephrased into 4j |  |  |  |  |
| f. Check for drug interactions of oral option with patient’s other medication | 5 | 91.67 | 4 | 70.83 | Accepted | Rephrased into 4k |  |  |  |  |
| g. Check for allergies to oral option | 5 | 91.67 | 4 | 70.83 | Accepted | Rephrased into 4l |  |  |  |  |
| h. Check patient adherence to oral option | 4.5 | 91.67 | 3 | 25.00 | Accepted | Rephrased into 4m |  |  |  |  |
| i. No vomiting within the last 24 hours |  |  |  |  |  | Result from rephrase 4d | 4 | 79.75 | Accepted | Accepted |
| j. Suitable oral switch option available |  |  |  |  |  | Result from rephrase 4e | 5 | 98.35 | Accepted | Rephrased into 4p |
| k. No contraindication or clinically significant drug interaction affecting oral switch option |  |  |  |  |  | Result from rephrase 4f | 5 | 95.45 | Accepted | Rephrased into 4p |
| l. No clinically significant allergy to oral switch option |  |  |  |  |  | Result from rephrase 4g | 5 | 96.69 | Accepted | Rephrased into 4p |
| m. No significant concerns over patient adherence to oral switch option |  |  |  |  |  | Result from rephrase 4h | 4 | 91.74 | Accepted | Rephrased into 4q |
| n. Gastrointestinal tract is functioning with no evidence of malabsorption |  |  |  |  |  |  |  |  |  | Result from rephrase 4a and 4c |
| o. Safe swallow or enteral tube administration |  |  |  |  |  |  |  |  |  | Result from rephrase 4b |
| p. Suitable oral switch option available, considering oral bioavailability, any clinically significant drug interactions or patient allergies |  |  |  |  |  |  |  |  |  | Result from rephrase 4j-m |
| q. No significant concerns over patient adherence to oral treatment |  |  |  |  |  |  |  |  |  | Result from rephrase 4m |
| 5. **Infection exceptions (at Step One) > Infection exclusions (at Step Three) > Special consideration (at Step Four)** | | | | | | | | | | |
| a. Deep-seated infections | 4 | 58.33 | 3 | 41.67 | Uncertain | Rephrased into 5p |  |  |  |  |
| b. Infections requiring high tissue concentration | 4 | 62.50 | 2 | 25.00 | Accepted | Rephrased into 5q |  |  |  |  |
| c. Infections requiring prolonged IV therapy | 4 | 70.83 | 3 | 33.33 | Accepted | Rephrased into 5r |  |  |  |  |
| d. Critical infection with high risk of mortality | 4 | 75.00 | 3 | 37.50 | Accepted | Accepted | 5 | 92.15 | Accepted | Accepted |
| e. On microbiology advice | 4 | 66.67 | 4 | 62.50 | Accepted | Rejected |  |  |  |  |
| f. Endocarditis | 4 | 75.00 | 3 | 45.83 | Accepted | Accepted | 5 | 87.19 | Accepted | Accepted |
| g. Meningitis | 4.5 | 83.33 | 3 | 45.83 | Accepted | Accepted | 5 | 89.26 | Accepted | Accepted |
| h. Bacteraemia, including *Staph. aureus* | 4 | 75.00 | 4 | 54.17 | Accepted | Accepted | 4.5 | 86.36 | Accepted | Rephrased into 5t |
| i. Immunocompromised | 3.5 | 50.00 | 4 | 62.50 | Rejected | Rejected |  |  |  |  |
| j. Abscess | 3 | 41.67 | 3 | 45.83 | Rephrased to ‘Undrained abscess’ | Rephrased into 5s |  |  |  |  |
| k. Severe or necrotising soft tissue infections | 4 | 70.83 | 3 | 41.67 | Accepted | Accepted | 5 | 90.08 | Accepted | Accepted |
| l. Infections of foreign bodies | 3.5 | 50.00 | 3 | 33.33 | Rejected | Rejected |  |  |  |  |
| m. Osteomyelitis | 4 | 54.17 | 2 | 33.33 | Uncertain | Accepted | 4 | 77.27 | Accepted | Accepted |
| n. Septic arthritis | 4 | 58.33 | 3 | 33.33 | Uncertain | Accepted | 4 | 79.34 | Accepted | Accepted |
| o. Empyema | 4 | 54.17 | 3 | 37.50 | Uncertain | Accepted | 4 | 71.07 | Accepted | Accepted |
| p. Deep-seated infection |  |  |  |  |  | Result from rephrase 5a | 4 | 81.40 | Accepted | Accepted |
| q. Infection requiring high tissue concentration |  |  |  |  |  | Result from rephrase 5b | 4 | 78.93 | Accepted | Accepted |
| r. Infection requiring prolonged IV therapy |  |  |  |  |  | Result from rephrase 5c | 5 | 89.26 | Accepted | Accepted |
| s. Undrained abscess |  |  |  |  |  | Result from rephrase 5j | 4 | 69.83 | Accepted | Accepted |
| t. Bloodstream infection |  |  |  |  |  |  |  |  |  | Result from rephrase 5h |
