## Supplementary material for "A Delphi Process for Reaching Nationwide Consensus on Antimicrobial Intravenous-to-oral Switch Criteria and Development of an Antimicrobial Intravenous-to-oral Switch Decision Aid": Pre Delphi_IVOS Criteria

**Table SX.** Forty-two intravenous-to-oral switch (IVOS) criteria from rapid review of the published literature (Literature), informed by hospital IVOS policies (Hospital) and expert advice (Expert) taken forward into the 4-step Delphi process.

| IVOS criteria | Source of criteria | Number of Literature papers (n=16) and  Trust policies (n=45) (%) | Evidence rating (if Literature) where H=High, M=Medium |
| --- | --- | --- | --- |
| 1. Timing of IV antimicrobial review | | | |
| a. Review antimicrobial within 24 hours | Hospital | 11 (24) |  |
| b. Review antimicrobial within 24-48 hours | Hospital | 5 (11) |  |
| c. Review antimicrobial within 48 hours | Literature  Hospital | 1 (6)  13 (29) | [H] |
| d. Review antimicrobial within 48-72 hours | Literature  Hospital | 3 (19)  5 (11) | [M] x2, [H] |
| 2. Clinical signs and symptoms | | | |
| a. Clinical signs and symptoms should be improving | Literature  Hospital | 9 (56)  29 (64) | [M] x8, [H] |
| 3. Infection markers | | | |
| a. Temperature should be between 36-38°C | Literature  Hospital | 2 (13)  4 (9) | [M] x2 |
| b. Temperature should be between 36-38 °C past 24 hours | Literature  Hospital | 3 (19)  14 (31) | [M] x2, [H] |
| c. Heart rate should be below 90 beats per minute | Literature  Hospital | 1 (6)  13 (29) | [H] |
| d. Heart rate should be below 90 beats per minute for past 12 hours | Literature | 1 (6) | [M] |
| e. Heart rate should be below 90 beats per minute for past 24 hours | Hospital | 4 (9) |  |
| f. Blood pressure should be stable | Literature | 1 (6) | [M] |
| g. Blood pressure stable for past 24 hours | Literature  Hospital | 2 (13)  4 (9) | [M] x2 |
| h. Respiratory rate should be below 20 breaths per minute | Literature  Hospital | 2 (13)  14 (31) | [M], [H] |
| i. Respiratory rate should be below 20 breaths per minute for past 24 hours | Literature  Hospital | 1 (6)  6 (13) | [M] |
| j. White cell count should be normalising | Literature  Hospital | 3 (19)  20 (44) | [M] x2, [H] |
| k. White cell count should be between 4 and 12 x10^9/L | Literature  Hospital | 3 (19)  6 (13) | [M] x2, [H] |
| l. White cell count should be between 4 and 12 x10^9/L or normalising | Literature  Hospital | 1 (6)  12 (27) | [M] |
| m. C-reactive protein should be normalising | Literature  Hospital | 2 (13)  15 (38) | [M] x2 |
| n. C-reactive protein does not reflect severity of illness or the need for IV antibiotics, and may remain elevated as the infection improves | Hospital | 17 (38) |  |
| 4. Enteral route | | | |
| a. Gastrointestinal tract must be functional | Literature | 9 (57) | [M] x8, H |
| b. Patient can tolerate/swallow oral option | Literature  Hospital | 9 (57)  39 (87) | [M] x6, [H]x3 |
| c. No evidence of malabsorption | Literature  Hospital | 11 (69)  30 (67) | [M] x9, [H] x2 |
| d. No vomiting | Literature  Hospital | 5 (31)  16 (35) | [M] x5 |
| e. There should be a suitable oral option available | Hospital | 28 (62) |  |
| f. Check for drug interactions of oral option with patient’s other medication | Hospital | 4 (9) |  |
| g. Check for allergies to oral option | Hospital | 5 (11) |  |
| h. Check patient adherence to oral option | Expert |  |  |
| 5. Infection exclusions | | | |
| a. Deep-seated infections | Literature | 3 (19) | [M] x3 |
| b. Infections requiring high tissue concentration | Literature | 3 (19) | [M] x2, [H] |
| c. Infections requiring prolonged IV therapy | Literature | 4 (25) | [M] x3, [H] |
| d. Critical infection with high risk of mortality | Literature | 1 (6) | [M] |
| e. On microbiology advice | Expert |  |  |
| f. Endocarditis | Literature  Hospital | 12 (75)  42 (95) | [M] x9, [H] x3 |
| g. Meningitis | Literature  Hospital | 9 (56)  41 (91) | [M] x7, [H] x2 |
| h. Bacteraemia, including *Staph. aureus* | Literature  Hospital | 9 (56)  43 (76) | [M] x9 |
| i. Immunocompromised | Literature  Hospital | 3 (19)  33 (73) | [M] x2, [H] |
| j. Abscess | Literature  Hospital | 7 (44)  31 (69) | [M] x7 |
| k. Severe or necrotising soft tissue infections | Literature  Hospital | 5 (31)  29 (64) | [M] x5 |
| l. Infections of foreign bodies | Literature  Hospital | 6 (38)  29 (64) | [M] x5, [H] |
| m. Osteomyelitis | Literature  Hospital | 6 (38)  29 (64) | [M] x5, [H] |
| n. Septic arthritis | Literature  Hospital | 5 (31)  28 (62) | [M] x5 |
| o. Empyema | Literature  Hospital | 5 (31)  23 (51) | [M] x5 |
